# Isoform-level analyses of 6 cancers uncover extensive genetic risk mechanisms undetected at the gene-level

**DOI:** 10.1101/2024.10.29.24316388

**Authors:** Yung-Han Chang, Sean T. Bresnahan, S. Taylor Head, Tabitha Harrison, Yao Yu, Chad D. Huff, Bogdan Pasaniuc, Sara Lindström, Arjun Bhattacharya

**Affiliations:** Quantitative Sciences Program, The University of Texas MD Anderson Cancer Center UTHealth Houston Graduate School of Biomedical Sciences, Houston, TX, USA; Department of Epidemiology, University of Texas MD Anderson Cancer Center, Houston, TX, USA; Department of Epidemiology, School of Public Health, University of Washington, Seattle, WA, USA; Department of Genetics, Perelman School of Medicine, University of Pennsylvania, Philadelphia, PA, USA; Public Health Sciences Division, Fred Hutchinson Cancer Center, Seattle, WA, USA; Institute for Data Science in Oncology, University of Texas MD Anderson Cancer Center, Houston, TX, USA

## Abstract

Integrating genome-wide association study (GWAS) and transcriptomic datasets can help identify mediators for genetic risk of cancer. Traditional methods often are insufficient as they rely on total gene expression measures and overlook alternative splicing, which generates different transcript-isoforms with potentially distinct effects. Here, we integrate multi-tissue isoform expression data from the Genotype Tissue-Expression Project with GWAS summary statistics (all N > 20,000 cases) to identify isoform- and gene-level associations with six cancers (breast, endometrial, colorectal, lung, ovarian, prostate) and six related cancer subtype classifications (N = 12 total). Directly modeling isoforms using transcriptome-wide association studies (isoTWAS) significantly improves discovery of genetic associations compared to gene-level approaches, identifying 164% more significant associations (6,163 vs. 2,336) with isoTWAS-prioritized genes enriched 4-fold for evolutionarily-constrained genes. isoTWAS tags transcriptomic associations at 52% more independent GWAS loci across the six cancers. Isoform expression mediates an estimated 63% greater proportion of cancer risk SNP heritability compared to gene expression. We highlight several isoTWAS associations that demonstrate GWAS colocalization at the isoform level but not at the gene level, including *CLPTM1L* (lung cancer), *LAMC1* (colorectal), and *BABAM1* (breast). These results underscore the importance of modeling isoforms to maximize discovery of genetic risk mechanisms for cancers.

## INTRODUCTION

Over the past twenty years, genome-wide association studies (GWASs) have successfully linked hundreds of genetic variants associated with increased risks for common cancers, including breast, prostate, lung, and colorectal cancers^1^. For example, Zhang and Ahearn et al. identified 32 new loci associated with breast cancer risk in European ancestry individuals^2^, totaling over 200 loci identified for overall and subtype-specific breast cancer risk. Similarly, Wang and Shen et al. identified 187 novel risk associations in a multi-ancestry prostate cancer GWAS^3^. Despite these findings underscoring the potential of GWASs to uncover genetic risk factors for cancer, a significant challenge remains as most genome-wide significant variants are in non-coding regions of the human genome, making it difficult to understand the biological mechanisms underlying these genetic associations.

To address this gap, transcriptome-wide association studies (TWASs) have emerged as a powerful complementary approach, providing valuable insights into the functional impact of genetic variants^4–8^. By focusing on the regulatory effects of genetic variants on gene expression, TWASs can identify potential target genes and pathways involved in disease processes. This integrated approach enhances our ability to translate genetic findings from GWASs into a deeper molecular understanding of disease etiology, which is necessary for improved diagnostics, prevention strategies, and therapeutic interventions in oncology.

Despite the promise of TWASs and related approaches, recent work suggests that gene expression may be a poor mediator of genetic risk of complex traits, potentially owing to differences in evolutionary pressures on genetic variants with large effects on complex traits and those with large effects on molecular phenotypes^9,10^. It is possible that part of this poor mediation is due to an overreliance on total gene expression as a fundamental unit of measure of the transcriptome. Total gene expression does not account for alternative splicing that can produce multiple isoforms from the same gene, which are often under subtle genetic or environmental control. The mRNA and protein isoforms generated through alternative processing of primary RNA transcripts can vary in function and potentially affect cancer risk in different ways^11,12^. To address this limitation, we applied isoform-level TWAS (isoTWAS), a framework that integrates genetic and isoform-level transcriptomic variation with GWAS summary statistics. This approach has previously demonstrated strong performance in increasing the discovery of brain-relevant traits, identifying transcriptomic association at far more GWAS loci compared to traditional gene-level TWAS while controlling the false discovery rate^13,14^.

In this study, we utilized isoTWAS to assess the association between isoform expression and risk of six cancer types (breast^15,16^, endometrial^17^, colorectal^18^, lung^19^, ovarian^20^, and prostate^21^), and their subtype classifications where applicable. We used publicly available European-ancestry cancer GWAS summary statistics and multi-tissue isoform- and gene-level expression data from the Genotype Tissue-Expression Project (GTEx, N > 100)^22^. isoTWAS reveals more associated loci than traditional TWAS, highlighting its potential to uncover novel genetic insights and enhance our understanding of cancer biology.

## RESULTS

We conducted isoform-(isoTWAS) and gene-level transcriptome-wide association studies (TWAS) for a total of 12 cancer outcomes: (1-3) breast (BRCA; overall, estrogen receptor (ER)+, ER-)^15,16^, (4) colorectal (CRC)^18^, (5) lung (LUNG; overall)^19^, (6) lung adenocarcinoma (LUAD)^19^, (7) lung squamous cell carcinoma (LUSC)^19^ (8-9) ovarian (OVCA; overall, serous)^20^, (10-11) prostate (PRCA; overall, advanced)^21^, and (12) endometrial (UCEC)^17^. We integrated GWAS summary statistics (N = 63,053-228,951) with multi-tissue expression QTL data from individuals without cancer collected post-mortem from the Genotype Tissue-Expression Project (GTEx, N = 108-574) (**Figure 1**). We selected these 12 cancer subtypes based on their large GWAS sample sizes, particularly with sufficient case numbers, which are essential for ensuring robust and reliable downstream analyses. GWAS and GTEx sample sizes and assignments of relevant tissues to cancer outcomes are provided in **Supplemental Tables 1-2**. See **Methods**, **Supplemental Methods, Supplemental Figure 1** for further details.

**Figure 1.**
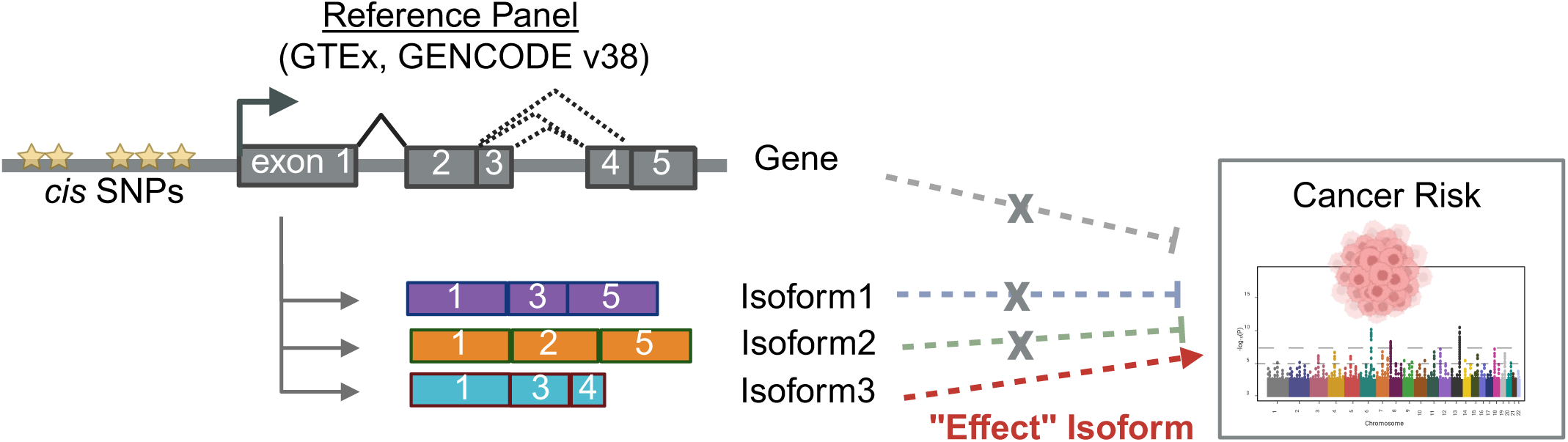
Importance of isoform expression analysis in cancer risk association. When examining only total gene expression, the gene shows no significant association with the cancer risk. However, analyzing expression at the isoform level reveals that a specific isoform of the gene is associated with the trait. This suggests that isoform-level mediation analysis can uncover regulatory mechanisms and prioritize genes that would otherwise be overlooked in total expression analyses.

### Isoform-level analysis reveals cancer risk associations not detected at the gene-level

In total, isoTWAS identifies 11,078 significant isoform associations across 6,163 unique genes, representing a ∼164% increase in gene identification as compared to traditional TWAS which identified 2,336 significant gene associations, (**Figure 2A; Supplemental Figure 2; Supplemental Table S3-5, Supplemental Data S1-2**). Of the genes identified, 10% from isoTWAS and 7% from traditional TWAS are included in the OncoKB Cancer Gene List, representing a moderate enrichment of hallmark cancer genes among isoTWAS gene (P = 0.03)^23,24^. For both TWAS and isoTWAS, most identified gene associations are observed for BRCA, ER+ BRCA, and PRCA, which also are the three largest GWAS. Specifically, isoTWAS identifies 3.12 times as many associations for BRCA (3,889 isoforms), 2.85 times more for PRCA (1,849 isoforms), and 2.39 times as many for ER+ BRCA (2,304 isoforms) as compared to TWAS, while showing relatively similar performance for LUNG; Manhattan plots of TWAS and isoTWAS results are shown in **Supplemental Figures S3-S14**. Given that the mean of the X^2^ distribution is linearly related to power and sample size, the percent increase in the test statistic serves as a measure of power or effective sample size. For X^2^ > 1, we calculated the percent increase for isoTWAS-based associations compared to TWAS-based associations (**Figure 2B**). Across 12 cancer outcomes, isoTWAS shows an average 25.3–37.4% increase in effective sample size compared to TWAS, suggesting that isoTWAS achieves comparable or greater power than TWAS, while requiring approximately 25–37% fewer samples (See **Methods** and **Supplemental Methods** for further details). Several key biological pathways are enriched for cancer-specific sets of isoTWAS-prioritized genes, including cell cycle and mitosis regulation, DNA and RNA binding, immune pathways, as well as downstream targets of cancer-relevant transcription factors like *ESR1*^25^, *RUNX2*^26^, and *YY1*^27^ (**Supplemental Table S6**, **Supplemental Figures S15-S20**).

**Figure 2:**
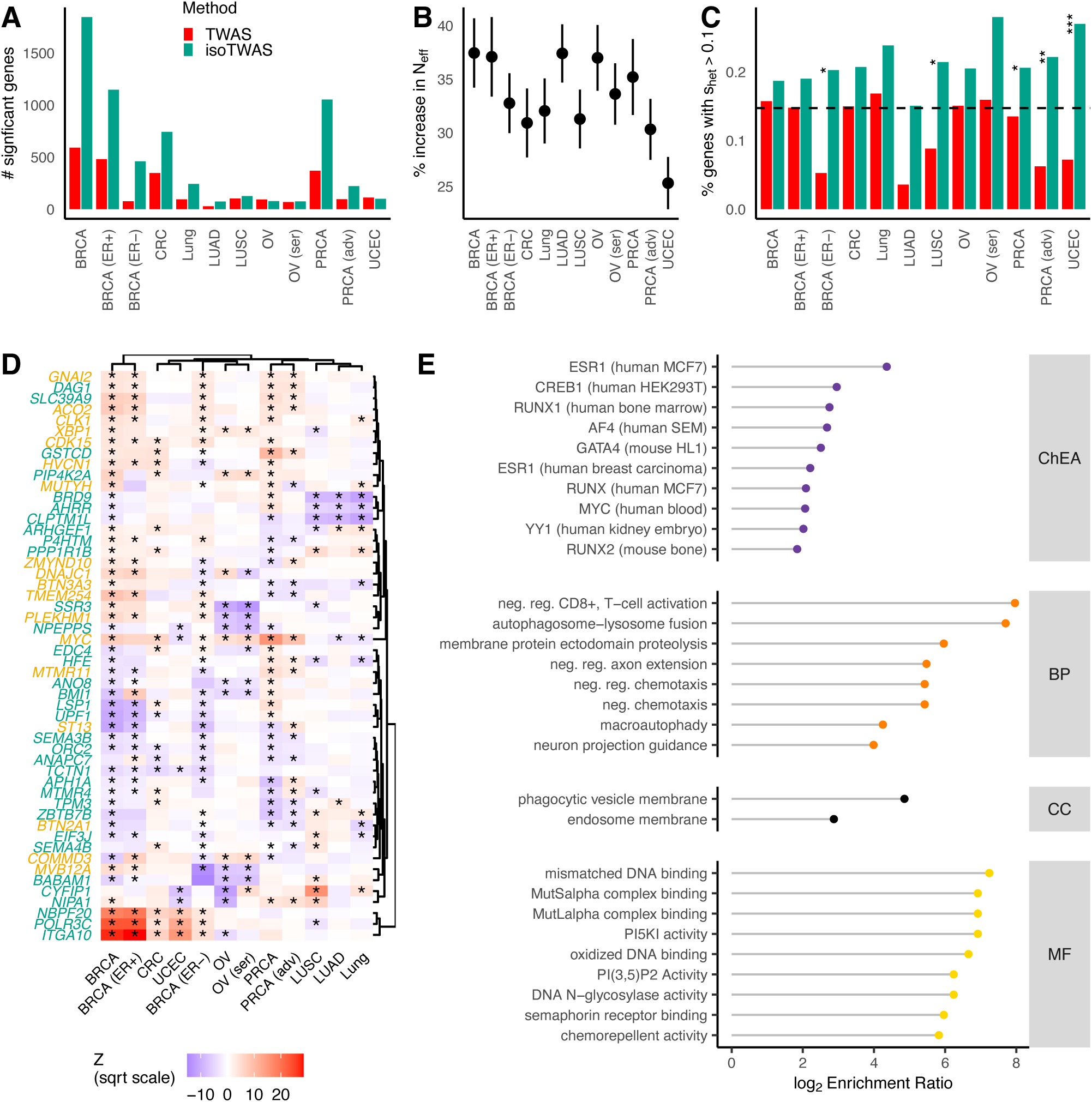
Isoform-level analysis identifies substantially more gene associations with cancer risk across 12 outcomes. **(A)** The number of unique transcriptome-wide significant genes identified with TWAS (red) and isoTWAS (green). **(B)** Percent increase in effective sample size with Wald-type 95% confidence interval using jackknife standard errors. **(C)** Proportion of transcriptome-wide significant genes identified with s_het_ > 0.1. Asterisks indicate FDR-adjusted P-value of X^2^ test for enrichment ratio of high pLI genes among all transcriptome-wide significant genes across method. (*) indicates FDR-adjusted P < 0.05, (**) P < 0.01, (***) P < 0.005. Black line shows the genome-wide proportion of genes with s_het_ > 0.1. **(D)** Scaled gene-level isoTWAS Z-score for 52 genes with isoform-level risk associations with at least 5 cancer outcomes across 12 outcomes. Genes are marked in green if no gene-level risk associations with any cancer outcome, and asterisk is shown if the gene association is significant for the given cancer outcome. **(E)** Log-enrichment ratio (X-axis) of over-represented gene ontologies (Y-axis), across ChIP-seq identified transcription factor targets (ChEA), biological process (BP), cellular component (CC), or molecular function (MF) pathways for 34 genes with isoform-level risk associations with at least 5 cancer outcomes and no TWAS associations.

We further explored if cancer risk genes identified by TWAS and isoTWAS capture true disease signals by identifying genes under selective constraint. If a gene is constrained, selection will act to remove variants that diminish gene function from the population, such as loss-of-function (LOF) variants. Here, we used Bayesian estimates of the s_het_ measure of constraint^28^, which, unlike traditional measures of constraint, is not biased towards longer genes. In total, 19.9% of isoTWAS gene associations (1,226 of 6,163) show s_het_ > 0.1, compared to 12.5% of TWAS gene association (293 of 2,336) with s_het_ > 0.1. Not only does this represent a significant enrichment of high s_het_ among isoTWAS-prioritized gene associations compared to TWAS-prioritized gene associations (X^2^ test P = 3.7 × 10^−15^), isoTWAS-prioritized genes are significantly enriched compared to the genome-wide proportion of high s_het_ genes (14.7%, X^2^ P < 2.2 × 10^−16^). In addition, we find significant enrichments of high s_het_ genes among transcriptome-wide significant genes for ER-BRCA, LUNG, LUSC, PRCA, Adv PRCA, and UCEC (X^2^ FDR-adjusted P-value < 0.05; **Figure 2C, Supplemental Table S4**).

Lastly, isoTWAS identifies 52 genes (34 undetected by TWAS) that are associated with five or more cancer outcomes (**Figure 2D**), including multiple known oncogenes or tumor suppressor genes, such as *MYC*^29^, *MUTYH*^30^*, GNAI2*^31,32^, *ACO2*^33^, and *BMI1*^34^. In comparison, TWAS identified 15 such genes, with four genes shared between the two methods. The 34 genes undetected by TWAS are enriched for multiple salient pathways: regulation of CD8-positive and T cells, regulation of membrane protein complexes, and phagocytic and autolysosome function. Additionally, these 34 genes are highly enriched for downstream targets of crucial oncogenic transcription factors, like *ESR1*, *GATA4*, *YY1,* and *MYC*, all of which were determined using ChIP-Seq experiments in human or mouse cancer cells or tumors (**Figure 2E**)^35^. We also observed that for many genes, the directions of isoform-level effects vary across traits (**Supplemental Figure 21**). Genes associated with a larger number of cancers tend to show greater expression variability across tissues. For many of these genes, such as *MYC*, distinct isoforms can show opposing effects (**Supplemental Figure 22**). Altogether, not only can isoTWAS identify constrained susceptibility genes for multiple cancers, but it can also reveal pan-cancer risk signals that TWAS is unable to detect, possibly due to opposing directions of effects of two or more isoforms of the same gene.

### Isoform expression explains more GWAS loci and overall SNP heritability of cancer risk

In addition to increasing the discovery of susceptibility genes across the genome, isoTWAS can identify far more transcriptomic mechanisms within independent, high-confidence genome-wide significant loci. Across all cancer outcomes, we identify 622 risk-associated GWAS SNPs (P < 5 × 10^−8^) in independent linkage disequilibrium (LD) blocks (see **Methods**). Among these 622 loci, 288 (46.3%) are tagged by TWAS, and 439 (70.6%) are tagged by isoTWAS, with 249 (40.0%) loci identified by both TWAS and isoTWAS (**Figure 3A, Supplemental Table S7**). In total, this represents an increase in significant associations in GWAS loci of 52.4% when using isoTWAS, rather than TWAS. Additionally, isoTWAS identified 2,911 unique significant genes outside of known GWAS loci.

**Figure 3:**
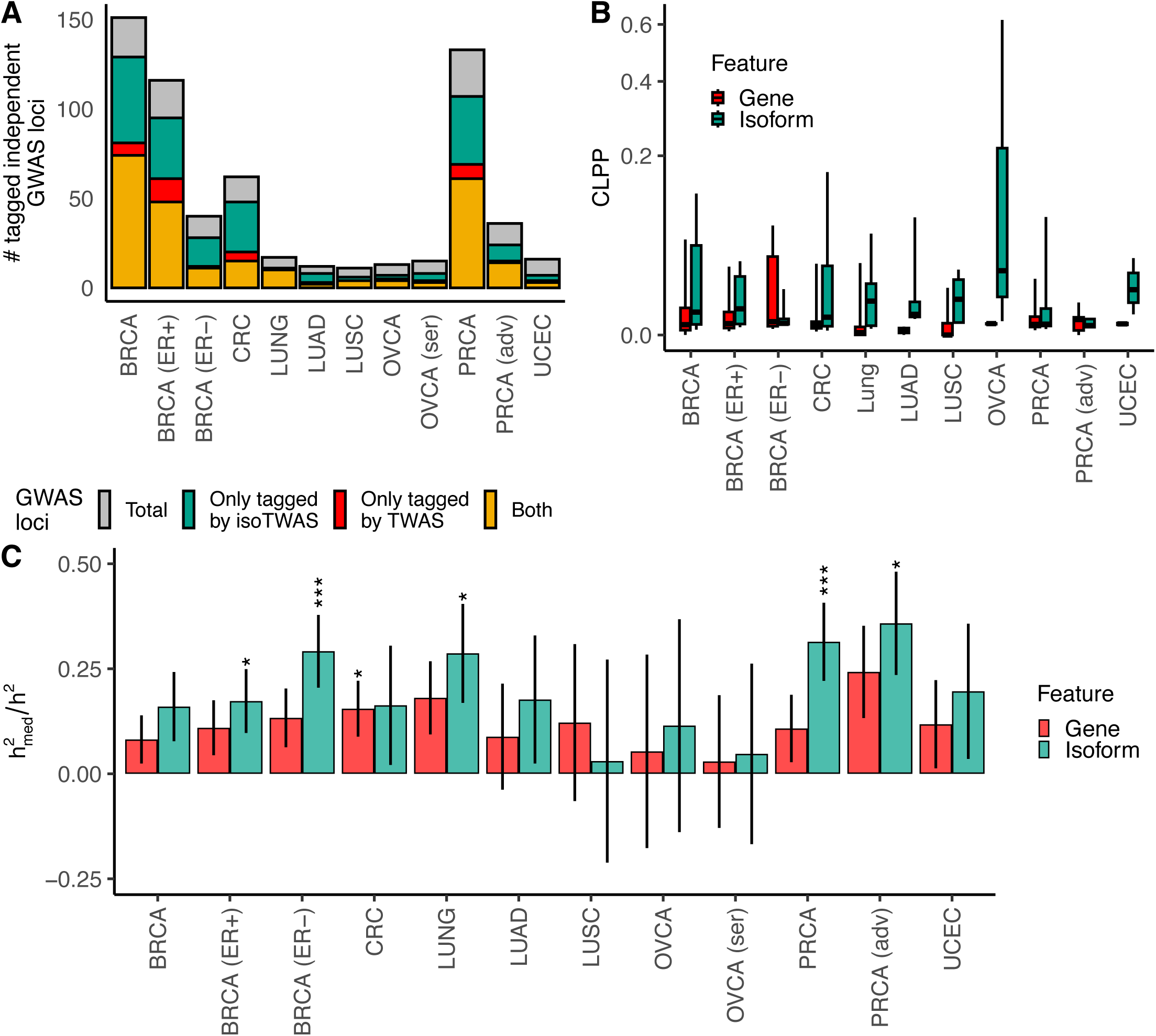
Isoform expression potentially mediates far more GWAS signal than gene expression. **(A)** The number of independent GWAS loci tagged by TWAS (red) or isoTWAS (green) or both in common (gold). **(B)** Boxplots of CoLocalization Posterior Probability (CLPP, Y-axis, square-root axis) of GWAS and gene expression QTL (red) and isoform expression QTL (green) for genomic loci with a GWAS P < 5×10^−8^ and QTL P < 10^−6^. **(C)** Ratio of gene-(red) and isoform-level (green) expression mediated heritability (h^2^_med_) and total SNP heritability (h^2^) with standard errors. Asterisks indicate FDR-adjusted P-value of Wald-type Z tests of h^2^_med_/h^2^ = 0. (*) indicates FDR-adjusted P < 0.05, (**) P < 0.01, (***) P < 0.005.

We also considered an orthogonal analysis to compare GWAS follow-up using both gene and isoform expression. We mapped *cis*-expression quantitative trait loci (eQTLs) for gene and isoform expression and conducted Bayesian colocalization analysis with GWAS signals using eCAVIAR^36^ to estimate the CoLocalization Posterior Probability (CLPP; see **Methods; Supplemental Table S8, Supplemental Data S3**). Generally, eQTLs with GWAS loci (**Figure 3B**), with the proportion of loci having isoform expression Quantitative Trait Loci (isoQTL) colocalizations with CLPP > 0.01 consistently higher across the 12 cancer outcomes (**Supplemental Figure S23**). Notably, in OVCA and UCEC, isoTWAS shows significantly higher CLPPs (median = 0.026, P = 0.013) than TWAS (median = 0.001, P = 0.001), suggesting that isoTWAS more effectively captures colocalized signals in for these cancer outcomes. Additionally, we estimated the proportion of total SNP heritability (h^2^) mediated by gene- and isoform-level expression (h^2^_med_), (**Figure 3C** and **Supplemental Table S9)**. Overall, isoform-level expression explains 62.7% more of cancer risk SNP heritability (19.2 ± 10.3%) compared to gene-level expression (11.8 ± 5.7%). Wald-type tests reveal that isoform expression mediates a significant proportion of cancer h^2^, with gene expression only explaining a significant portion for ER-BRCA (isoTWAS: 0.291, FDR-adjusted P = 0.005; TWAS: 0.133, P = 0.068), ER+ BRCA (isoTWAS: 0.173, P = 0.045; TWAS: 0.110, P = 0.102), LUNG (isoTWAS: 0.287, P = 0.044; TWAS: 0.181, P = 0.056), PRCA (isoTWAS: 0.314, P = 0.005; TWAS: 0.108, P = 0.164), and Adv PRCA (isoTWAS: 0.358, P = 0.014; TWAS: 0.243, P = 0.046). These results indicate that not only does isoTWAS recapitulate an overwhelming majority of TWAS signals at GWAS loci, isoTWAS substantially increases discovery of candidate GWAS mechanisms and transcriptomic features that potentially mediate genetic effects on cancer risk.

### isoTWAS and isoform-eQTL colocalization prioritize undetected mechanisms at GWAS loci

A main goal of isoform-specific analyses is to nominate a more granular hypothesis of transcriptomic regulation. Post-hoc transcript-level fine-mapping reveals nine loci where isoTWAS prioritization coincides with isoform-eQTL colocalization (CLPP > 0.01) but no gene-eQTL colocalization (**Methods**; **Supplemental Data S3**), across BRCA (**Figure 4**, **Supplemental Figures S24**), ER-BRCA (**Supplemental Figure S25-S26**), CRC (**Figure 5**, **Supplemental Figure S27**), LUNG (**Figure 6**), PRCA (**Supplemental Figure S28**), and UCEC (**Supplemental Figure S29**). We highlight *CLPTM1L*, *LAMC1,* and *BABAM1* here due to previously-reported pleiotropic associations across multiple cancers. *CLPTM1L* is located near the *TERT* locus, known for its pleiotropic links to various cancers, including ER-BRCA, CRC, glioma, LUNG, melanoma, OVCR, pancreatic, and PRCA cancer ^37^, the *LAMC1* locus has shown associations with UCEC, glioma, and PRCA^38–40^, and *BABAM1* is a recognized GWAS hit with broad implications for cancer risk, including its interaction with the well-known breast cancer-related gene *BRCA1*^41–43^. Additional examples which follow a similar pattern are presented in the **Supplemental Results**. We leveraged (1) functional annotations from ENCODE and ROADMAP Epigenomic Project (**Supplemental Table 10)**, (2) splice-site QTL (sQTL) and 3’UTR alternative polyadenylation QTL (apaQTL) summary statistics from GTEx, and (3) predicted splicing potential of variants using SpliceAI^44^ to investigate chromatin state and regulatory activity or more detailed splicing mechanisms near the implicated loci^22,45–48^. For these examples, we refer to the isoform of a gene that passes all isoTWAS post-hoc analyses as the “prioritized isoform” and the top identified isoQTL by P-value as the isoform’s “lead isoQTL”.

**Figure 4:**
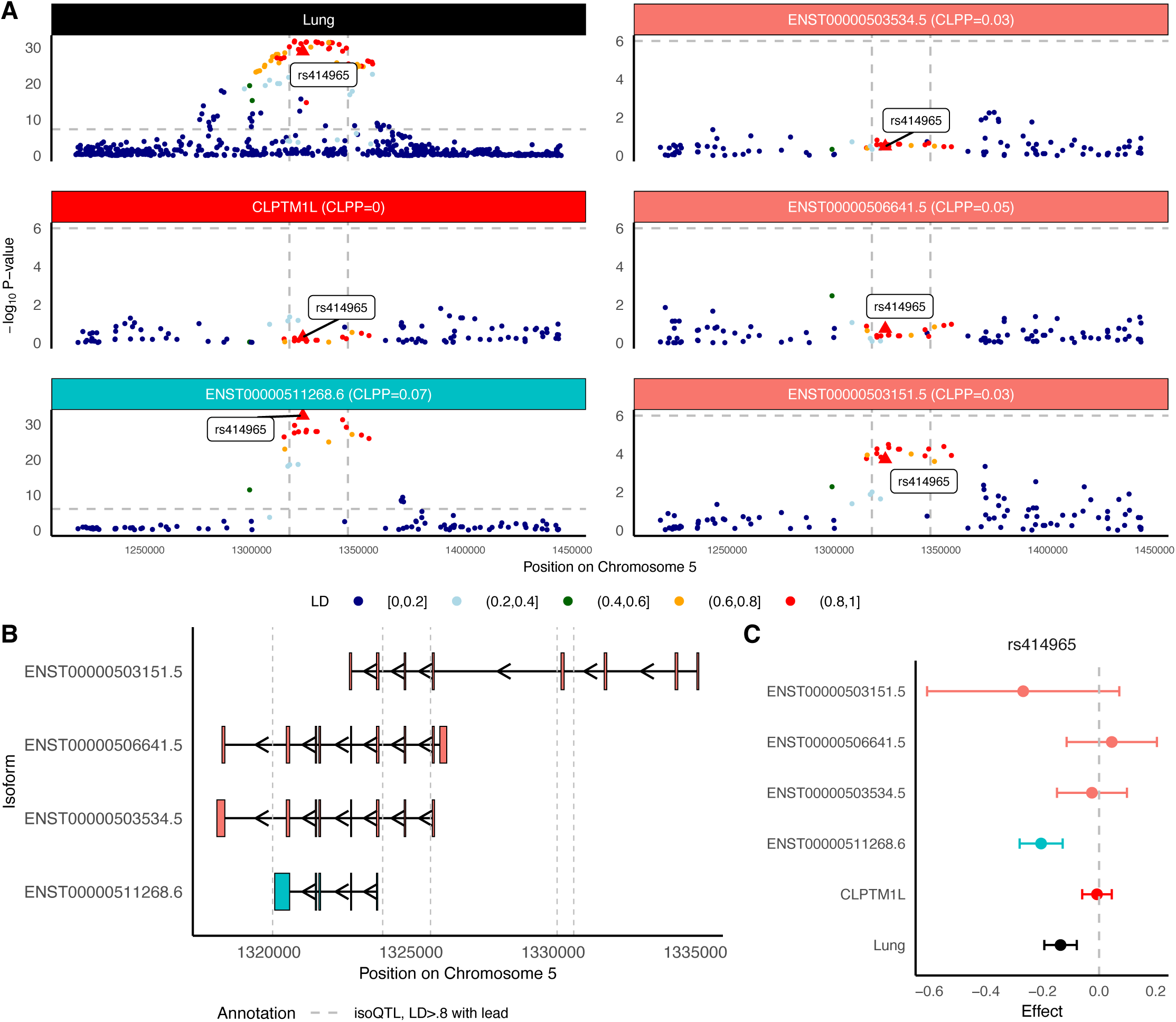
CLPTM1L isoforms may mediate lung cancer risk GWAS locus at Chromosome 5p15.33. **(A)** Manhattan plot of GWAS effects, *CLPTM1L* gene-eQTLs, and isoform-eQTLs for all significantly associated isoforms of *CLPTM1L*, colored by LD to rs414965, the lead isoQTL for ENST00000511268.6. **(B)** Transcript structure of significantly associated isoforms of *CLPTM1L*. Vertical lines indicated significant isoQTLs of LD > 0.8 to rs414965, strongest isoQTL of *CLPTM1L* isoforms. **(C)** SNP effect sizes on lung cancer risk (black), *CLPTM1L* gene expression (red), ENST00000511268.6 isoform expression (blue), and other expression of other isoforms (peach) for rs414965.

**Figure 5:**
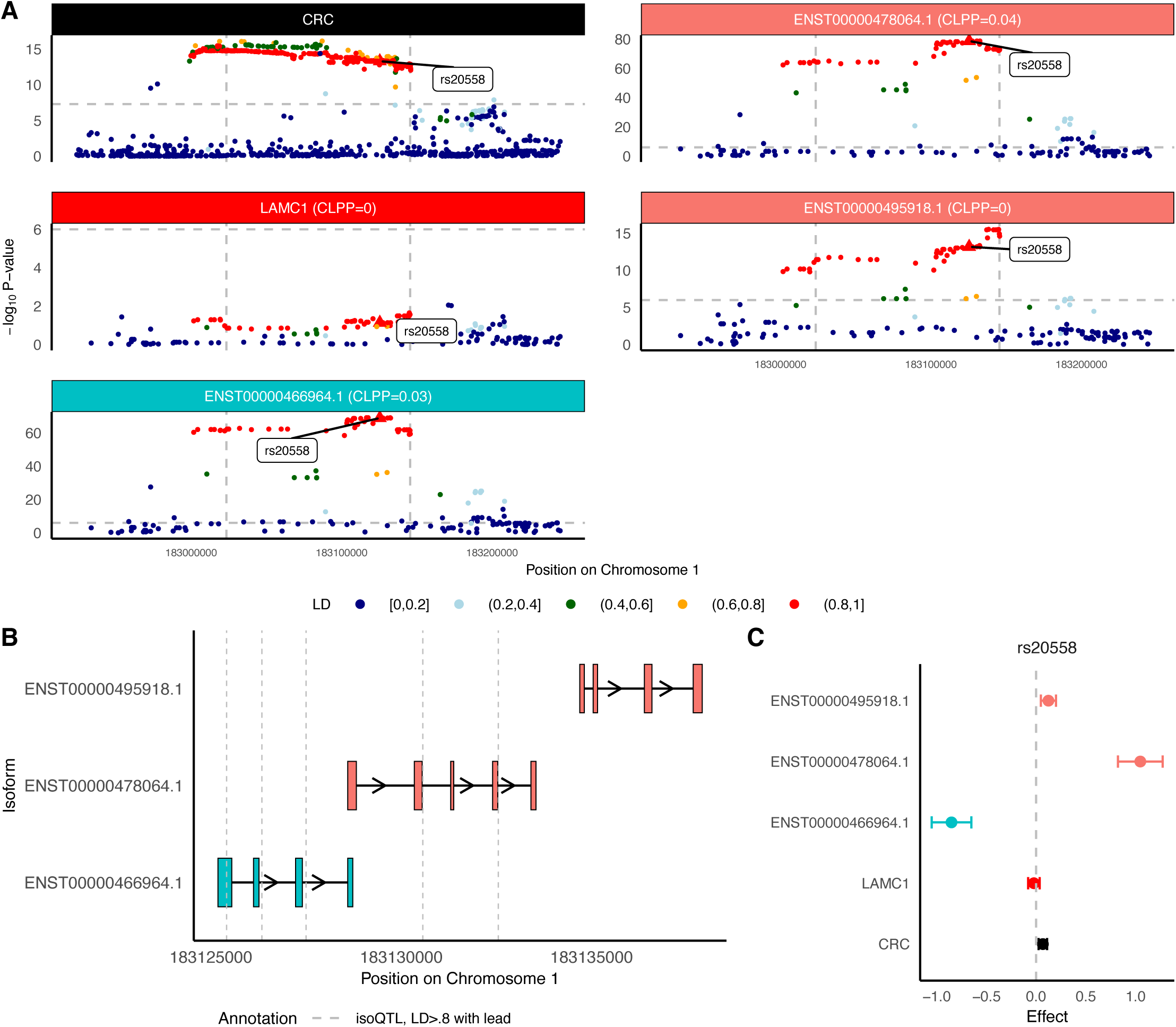
LAMC1 isoforms may mediate colorectal cancer GWAS locus at Chromosome 1q25.3. **(A)** Manhattan plot of GWAS effects, *LAMC1* gene-eQTLs, and isoform-eQTLs for all significantly associated isoforms of *LAMC1*, colored by LD to rs20558, the lead isoQTL for ENST00000466964.1. **(B)** Transcript structure of significantly associated isoforms of *BABAM1*. Vertical lines indicated significant isoQTLs of LD > 0.8 to rs20558, strongest isoQTL of *LAMC1* isoforms. **(C)** SNP effect sizes on colorectal cancer risk (black), *LAMC1* gene expression (red), ENST00000466964.1 isoform expression (blue), and other expression of other isoforms (peach) for rs20558.

**Figure 6:**
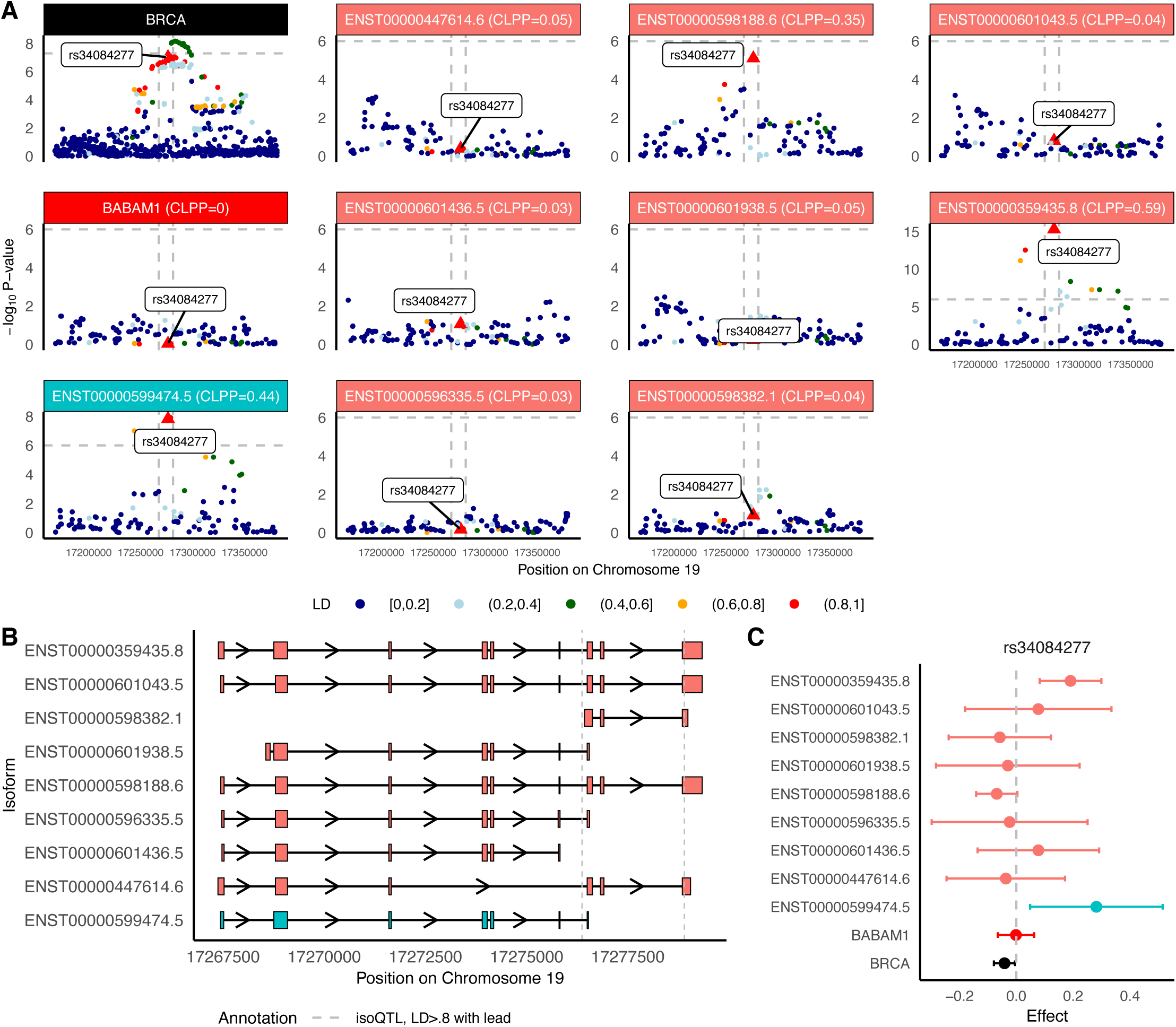
BABAM1 isoforms may mediate breast cancer risk GWAS locus at Chromosome 19p13.11. **(A)** Manhattan plot of GWAS effects, *BABAM1* gene-eQTLs, and isoform-eQTLs for isoforms of *BABAM1*, either prioritized through isoTWAS or with an isoQTL with P < 1e-6. **(B)** Transcript structure of *BABAM1*. Vertical lines indicated significant isoQTLs of LD > 0.8 to rs34084277, strongest isoQTL of *BABAM1* isoforms. **(C)** SNP effect sizes on breast cancer risk (black), *BABAM1* gene expression (red), expression of isoTWAS-prioritized isoforms (blue), and expression of other isoforms (peach) for rs34084277 (lead isoQTL).

First, four isoforms of *CLPTM1L* (Chromosome 5p15.33, s_het_ = 0.02, 16 total isoforms in GENCODE) are significantly associated with LUNG. We also find that isoforms of *CLPTM1L* are associated with BRCA, PRCA, LUAD, and LUSC. Fine-mapping prioritizes ENST00000511268.6 in the 90% credible set with Posterior Inclusion Probability (PIP) = 0.90. Although there are many genome-wide significant SNPs within the gene body of *CLPTM1L*, no gene-eQTL signal is observed (P < 10^−6^), whereas a strong ENST00000511268.6 isoform-eQTL signal colocalizes with the GWAS signal (CLPP = 0.07). Additionally, there is no strong isoform-eQTL signal for the other three isoforms identified via isoTWAS in this region. The lead isoQTL rs414965 is in high LD with multiple genome-wide significant SNPs (**Figure 4A**). The exons comprising ENST00000511268.6 overlap with those of ENST00000503534.5 and ENST00000506641.5 with significant isoform-eQTLs flanking ENST00000511268.6 (**Figure 4B**). However, rs414965 does not have significant effects on ENST00000503534.5, ENST00000506641.5, or ENST00000503151.5 expression (**Figure 4C**), despite its strong negative associations with LUNG and ENST00000511268.6 expression. We observed a CCCTC-binding factor (CTCF) peak at the terminal exon of the prioritized isoform ENST00000511268.6 and POLR2AphosphS5 in the last exon. Additionally, we find an alternative enhancer at the 5’ end of the prioritized isoform (**Supplemental Figure 30**). ENST00000511268.6 showed strong expression in lung, spleen, and small intestine, with limited differential isoform usage across tissues, and a strong negative correlation with ENST00000503534.5 (**Supplemental Figure 31**).

Next, three isoforms of *LAMC1* (Chromosome 1q25.3, s_het_ = 0.10, six total isoforms in GENCODE) are associated with CRC. Fine-mapping prioritizes only ENST00000466964.1 in the 90% credible set with PIP = 1. Again, genome-wide significant CRC-associated SNPs within the gene body show no gene-eQTL signal (P < 10^−6^) but a strong ENST00000466964.1 isoform-eQTL signal that colocalizes with the GWAS signal (CLPP = 0.03) is observed. For the other two isoforms, ENST00000478064.1 and ENST00000495918.1, the lead isoQTL rs20558 is in high LD with multiple genome-wide significant SNPs (**Figure 5A**). The exons of these three isoforms are generally distinct sets with significant isoform-eQTLs generally falling on the 3’ end of multiple exons (**Figure 5B**). rs20558 shows a significant association with increased CRC risk, no effect on *LAMC1* gene expression, and a significant decreasing effect on ENST00000466964.1 expression (**Figure 5C**). Interestingly, this same SNP has a significant increasing effect on the expression of ENST00000478064.1 and ENST00000495918.1, though only ENST00000478064.1 colocalizes with CRC risk at CLPP > 0.01. One of the SNPs in perfect LD (LD = 1) with our lead isoQTL is not present in the GTEx panel we used but shows potential splicing effects based on SpliceAI predictions^44,49^. rs34133998 may disrupt an existing donor site with stronger probability (Donor Loss = 0.77). The result supports a possible regulatory mechanism involving splicing (**Supplemental Table 11).** We observed splicing events that are associated with our perfect LD SNPs at the start site of the prioritized isoform ENST00000466964.1 in both subcutaneous adipose and colon tissues. We also observed an enhancer overlapping the first three exons of the prioritized isoform. The presence of POLR2A and POLR2AphosphoS5 peaks at the terminal exon provides supporting evidence for transcriptional pausing and termination at that site (**Supplemental Figure 32**). We identified a relatively higher expression in two tissues—subcutaneous adipose and cultured fibroblasts—at both the gene and transcript levels, while ENST00000495918.1 exhibited high isoform usage in whole blood tissue. (**Supplemental Figure 33**).

Lastly, nine isoforms of *BABAM1* (Chromosome 19p13.11, s_het_ = 0.05, 18 total isoforms in GENCODE) are associated with BRCA. In our study, isoforms of *BABAM1* are also associated with OVCA, OVCA ser, LUSC, and ER-BRCA in isoTWAS. Fine-mapping prioritizes ENST00000599474.5 in the 90% credible set and a PIP of 1. There are multiple genome-wide significant BRCA-associated SNPs (P < 5×10^−8^) within the *BABAM1* gene body showing no gene-eQTL signal (P < 10^−6^). Only ENST00000599474.5 (CLPP = 0.44) and ENST00000359435.8 (CLPP=0.59) show strong isoQTL effects that colocalize with the GWAS signal. In addition, the lead isoQTL rs34084277 for ENST00000599474.5 is in high LD (LD > 0.8) with multiple genome-wide significant SNPs (**Figure 6A**). We observed two splicing events associated with the SNPs in perfect LD with the lead isoQTL in both subcutaneous adipose and breast tissue. We also observed CTCF at the terminal exon (**Supplemental Figure 34**). We did not observe large variance in the expression of the prioritized isoform ENST00000599474.5 across tissues. ENST00000598188.5 exhibited high isoform usage; however, none of the transcripts demonstrated clear tissue-specific differences in isoform usage. Additionally, we found a high positive correlation among six transcripts, including the prioritized isoform (**Supplemental Figure 35**). The exon structure of these nine isoforms reveals a group of exons at the 3’ end of the gene body, which is flanked by the lead isoQTLs of ENST00000599474.5 (**Figure 6B**). Specifically, due to this unique exon structure and previous evidence of *BABAM1* harboring rare variants associated with BRCA risk^41–43^, we followed up on this locus using a rare-variant analysis in UK Biobank whole exome sequencing data^50^ using the Variant Annotation, Analysis, and Search Tool (VAAST2)^51,52^. We find three BRCA-associated isoforms in *BABAM1* enriched for risk-associated rare variants (MAF < 0.5%), with associations mainly concentrated in exons at the 5’ end and first exon at the 3’ end of the transcript (**Supplemental Methods; Supplemental Figure 36; Supplemental Tables S12-S13**). See **Supplemental Results** for further details. Lastly, rs34084277 has a strong protective effect on *BRCA* risk, no effect on *BABAM1* gene expression, and a significantly increasing effect on expression of ENST00000599474.5 and ENST00000359435.8 (**Figure 6C**).

Of note, isoTWAS associations for *CLPTM1L*, *LAMC1*, and *BABAM1* were all prioritized using models trained in subcutaneous adipose tissue. To investigate why these genes are identified specifically in subcutaneous adipose, we conducted a GTEx QTL analysis for the prioritized transcripts to determine whether this is due to tissue-specific effects or simply a matter of statistical power. **Supplemental Figure 37** shows that, for the prioritized isoforms, the lead isoQTL and those SNPS in high LD also have large QTL effects in the corresponding tissue of origin, suggesting that these isoform effects may be present in other tissues but suffer from low statistical power in these tissues.

## DISCUSSION

We show that integrating cancer risk GWAS summary statistics with isoform-level transcriptomic variation can greatly increase discovery of susceptibility genes for six cancers and their subtype classifications. By using isoTWAS rather than traditional gene-level TWAS, we identify nearly 2.5-fold more associations. More saliently, isoTWAS-identified genes are significantly enriched for evolutionarily constrained genes (s_het_ > 0.1), which previously studies have shown to be likely to contain clinically-relevant *de novo* or rare variants and hypothesize to predict drug toxicity and characterize transcriptional regulation^10,53^. Isoform expression, rather than total gene expression, captures more risk-associated genetic variation and exhibits stronger colocalization with GWAS signals. Additionally, though isoform expression alone does not reconcile most of the SNP heritability of cancer risk, isoform expression is estimated to mediate nearly twice as much heritability as gene expression, and nearly three times of the SNP heritability signal in the case of PRCA. Most importantly, isoTWAS can find isoform associations at 249 of 288 GWAS loci tagged by TWAS-identified genes and uniquely tag 190 additional GWAS loci that cannot be contextualized via TWAS. In aggregate, these findings underscore the utility of considering the transcriptome on the transcript-isoform, rather than gene-level. By modeling a different quantification of the same steady-state RNA-seq datasets with sample sizes of only up to ∼800, isoTWAS increases discovery specifically at GWAS loci by ∼52% without additional sequencing costs.

Our isoform-level fine-mapping coupled with eQTL colocalization identifies nine gene candidates with GWAS risk SNPs within the gene body that colocalize with isoQTLs despite exhibiting no gene-eQTL signal at P < 10^−6^. We discuss three of these gene candidates. First, isoTWAS identifies an association between ENST00000511268.6, an isoform of *CLPTM1L* (16 total isoforms), and LUNG. Studies spanning back 15 years have identified multiple polymorphisms within the *CLPTM1L* gene body associated with LUNG, as well as pleiotropic associations with other malignancies^54–58^. In particular, an *in vitro* study provided evidence that *CLPTM1L* is oncogenic, specifically for Ras-driven lung cancers, in line with its effect on protecting tumor cells from genotoxic apoptosis^59^, and is a promising therapeutic target for therapy-resistant tumors^60^. The regulatory annotation region reveals a CTCF peak at the terminal exon and POLR2AphosphoS5 signal at the last exon of the prioritized isoform ENST00000511268.6, suggesting transcription termination through polymerase stalling and recruitment of the splicing machinery, respectively. An enhancer at the 5’ end the lead transcript, suggesting regulatory activity in this region that may influence isoform expression. Further molecular studies are needed to test the association between the enhancer and the isoform. In non-cancerous lung tissue, strong transcriptional activity overlaps with three transcripts of *CLPTM1L,* whereas in lung tumor, it is largely confined to the prioritized isoform, suggesting that genetic regulation of the isoTWAS-prioritized isoform is potentially cancer-specific. *CLPTM1L* (Chromosome 5p15.33) is also local to *TERT*, which harbors pleiotropic associations with multiple cancers^37^. Our study identifies associations between *CLPTM1L* and PRCA, consistent with previous findings of SNPs in the 5p15.33 region associated with PRCA^61,62^. However, while *TERT* is known for its pleiotropic effects, we observed only gene-level associations for *TERT* in our analyses, with no isoTWAS associations detected for its specific isoforms. Further work is needed to fully interrogate this pleiotropy and assess tissue-specific isoform expression for both *TERT* and *CLPTM1L*, especially since *TERT* shows low expression across multiple tissues in GTEx^22^.

We find a similar pattern for colorectal cancer and ENST00000466964.1, an isoform of *LAMC1* (six total isoforms), a gene that regulates cell adhesion, differentiation, migration, and signaling, and lies at a pleiotropic locus for CRC, PRCA^38^, and obesity risk^63^. A previous study incorporating splicing measures has prioritized *LAMC1* as a novel transcriptome-mediated CRC locus^64^. Additionally, rs34295433 in *LAMC1* was identified as a susceptibility SNP for PRCA in a Taiwanese population^38^. However, in our study, isoTWAS did not detect any associations between *LAMC1* and PRCA. *LAMC1* belongs to the laminin family of extracellular matrix proteins that are significantly involved in survival and proliferation of cancer cells, angiogenesis, migration and basement membrane breach by cancer cells, and metastatic events^65^. Computational analyses of laminin proteins have shown their prognostic ability in colorectal cancer progression, placing higher weights for *LAMC1* compared to other constituents of the family^66^. One SNP in perfect LD with the lead isoQTL exhibits potential splicing disruption according to SpliceAI predictions, supporting a regulatory mechanism involving isoform-specific splicing. We observed splicing events associated with these SNPs at the start site of the prioritized isoform ENST00000466964.1 in both subcutaneous adipose and colon tissues, suggesting their potential influence on splicing regulation. Enhancer activity spanning the 5’ end of the transcript further supports a regulatory role in this region. Additionally, the presence of POLR2A and POLR2AphosphoS5 peaks at the terminal exon indicates potential transcriptional pausing and isoform-level regulatory control.

Lastly, isoTWAS detects an association between BRCA and ENST00000599474.5, an isoform of *BABAM1* (18 total isoforms), a gene involved in checkpoint signaling, regulation of DNA repair, and mitosis. *BABAM1* has previously been implicated as a low-penetrance risk locus that interacts with *BRCA1* in both triple-negative breast cancer and ovarian cancer risk^41–43^. Consistent with previous studies, isoTWAS also identifies associations within this region for OVCA, OVCA ser, and ER-BRCA. Observed splicing events associated with high-LD SNPs in both subcutaneous adipose and breast tissues further suggest a potential regulatory role in splicing. Additionally, the presence of a CTCF binding peak at the terminal exon supports the hypothesis of transcriptional termination at this region, potentially contributing to isoform-specific expression regulation. Additionally, our VAAST analysis for *BABAM1* in UKBB indicates that a group of exons at the 3’ end of the gene harbor potentially disease-causing rare variants (MAF < 0.01, only in coding regions). The lead isoQTL for our isoTWAS-prioritized isoform of *BABAM1* (MAF > 0.01, both coding and non-coding region) is upstream of these exons at the 3’ end, potentially influencing the splicing patterns specifically at the 3’ end of the gene. Further research is required to investigate how these isoQTLs influence splicing regulation and their specific role in tumorigenesis and cancer development, but a methodological opportunity may lie in integrating splicing- and isoQTLs with rare variants to identify transcriptomic mechanisms for cancer risk. These results underscore that the added resolution provided with isoform expression can lead to more specific mechanistic hypotheses for cancer risk and inform follow-up with functional studies, both *in silico* and experimental.

We conclude with three limitations of our work. First, the complexity of the *BABAM1* transcript structure and the sheer number of *BABAM1* isoforms with strong isoQTL signals underscores a methodological opportunity. We applied the FOCUS framework which leverages a non-informative prior that may be insufficient^67^. Fine-mapping of isoforms is challenging due to horizontal pleiotropy of SNP-isoform effects shared across exons and strong LD patterns. This horizontal pleiotropy can reduce power and increase false-positive rates. Incorporating classes of isoforms with shared exon sets may lead to improved fine-mapping, thereby increasing coverage and resolution of credible sets of isoforms within a GWAS locus.

Second, we derived isoform-level quantifications using Salmon, a method constrained by the limitations of short-read RNA-seq where maximum-likelihood estimates are based on transcriptomic annotations, introducing some uncertainty. The limited diversity of tissue-specific annotated isoforms may affect the accuracy of these estimates. In contrast, long-read RNA-seq can capture splicing and structural variation missed by short-read RNA-seq, revealing more complex and rarer transcript-isoforms. As long-read RNA-seq becomes increasingly scalable, its comprehensive view of transcriptomes will allow for more precise quantification of isoforms potentially improving isoTWAS performance. Additionally, the prioritization of these three genes in subcutaneous adipose tissue may reflect tissue-specific regulatory mechanisms, but it could also result from the statistical power across tissues, driven by sample size or the robustness of isoform quantification. Our QTL analysis indicates that the isoQTL effects observed in adipose tissue are also detectable in the tissues of origin, and the regulatory architecture appears similar between adipose and the original tissue. This may suggest that the observed isoTWAS associations in subcutaneous adipose and not the tissue of origin are likely due to power limitations rather than true tissue specificity. Long-read sequencing technologies, which offer improved isoform-level resolution, may help clarify the robustly expressed isoforms in each tissue and reveal genuine tissue-specific genetic regulation at risk loci. Third, as current eQTL datasets are mostly generated in populations of European ancestry, we focused the current analyses on cancer GWAS summary statistics based on individuals of European ancestry, as expression models trained in predominantly European-ancestry cohorts fail to predict expression accurately in individuals of different ancestries^68–71^. Future data generation should increase diversity in molecular datasets accompanied by methodological research to model multi-ancestry samples across eQTL and GWAS datasets.

## METHODS

### Data collection

#### GTEx genotype and transcriptomic data

For 48 tissues from GTEx^22^, we quantified RNA-seq data (all N > 100) using Salmon v1.5.2 in mapping-based mode^72^. We built a decoy-aware transcriptomic index in Salmon with GENCODE v38 transcript sequences and the full GRCh38 reference genome as decoy sequences^45^. Salmon was then run on FASTQ files with mapping validation and corrections for sequencing and GC bias. We then imported Salmon isoform-level quantifications and aggregated to the gene-level using tximeta v1.16.1^73^. Using edgeR, gene and isoform-level quantifications underwent TMM-normalization, followed by transformation into a log-space using the variance-stabilizing transformation using DESeq2 v1.38.3^74,75^. We then residualized isoform-level and gene-level expression (as log-transformed CPM) by all tissue-specific covariates (clinical, demographic, genotype principal components (PCs), and expression PEER factors) used in the original QTL analyses in GTEx.

SNP genotype calls were derived from Whole Genome Sequencing data from individuals of European ancestry, filtering out SNPs with minor allele frequency (MAF) less than 5% or that deviated from HWE at *P* < 10^−5^. We further filtered out SNPs with MAF less than 1% frequency among the European ancestry samples in 1000 Genomes Project^76^. Due to limited sample sizes in other ancestry groups in GTEx, we focused solely on European ancestry for this study.

### GWAS summary statistics

We obtained GWAS summary statistics for risk of 12 cancer outcomes, all from samples of European-ancestry individuals^77^: overall breast cancer (BRCA; 122,977 cases/105,974 controls)^15^, estrogen-receptor positive breast cancer (ER+ BRCA; 69,501 cases/95,042 controls)^15^, estrogen-receptor negative breast cancer (ER-BRCA; 30,882 cases/110,058 controls)^16^, Colorectal cancer (CRC; 55,168 cases/65,160 controls)^18^, overall lung cancer (LUNG; 29,266 cases/56,450 controls)^19^, lung adenocarcinoma (LUAD; 11,273 cases/55,483 controls)^19^, lung squamous cell Carcinoma (LUSC; 7,426 cases/55,627 controls)^19^, overall ovarian cancer (OVCA; 22,406 cases/40,951 controls)^20^, serous ovarian cancer (OVCA ser; 19,890 cases/68,502 controls)^20^, overall prostate cancer (PRCA; 79,166 cases/61,106 controls)^21^, advanced prostate cancer (Adv PRCA; 15,167 cases/58,308 controls)^21^, endometrial cancer (UCEC; 12,906 cases/108,979 controls)^17^. We selected these 12 cancer subtypes based on their large GWAS sample sizes.

### Statistical analysis

#### Code and data availability

Sample scripts for analyses are available from github.com/bhattacharya-a-bt/MultiCancerIsoTWAS. Links to GWAS summary statistics are provided in **Supplemental Table S1**. isoTWAS and TWAS models are available from https://zenodo.org/records/11048201^78^. GTEx genotyping and expression data were accessible from dbGaP accession number phs000424.v9. Supplemental Data Tables S1-S3 can be downloaded from https://zenodo.org/records/14010391.

### Isoform- and gene-level transcriptome-wide association studies

We trained predictive models of gene and isoform expression using all *cis*-SNPs within 1 Mb of the gene body for the 12 cancer outcomes (**Supplemental Figure 1A**), using predictive models described previously^13^. We selected the best gene or isoform model through a 10-fold cross-validation and retained genes and isoforms that could be predicted at cross-validation R^2^ > 0.01. Using these predictive models, we employed a weighted burden test to estimate the association between the genetically-predicted component of a gene or isoform with cancer risk (**Supplemental Figure 1B**). To account for multiple testing burden for isoform associations, we used a two-stage testing framework, as described previously^13^ (**Supplemental Figure 1C**). At the end of these two steps, isoTWAS identified a set of genes and their isoforms that were associated with the trait.

For significant genes (from TWAS) and isoforms (from isoTWAS), we tested whether the SNP-gene/isoform effects from the predictive models add additional information beyond the SNP-risk effects from the GWAS through a conservative permutation test. We permuted the SNP-gene/isoform effects in the predictive models 10,000 times and generated a null distribution for the gene/isoform test statistic, which was used to calculate a P-value. We only conducted this permutation test for genes from TWAS with FDR-adjusted P < 0.05 and isoforms from isoTWAS with FDR-adjusted P < 0.05 and FWER-adjusted P < 0.05. After permutation testing, we conducted isoform- and gene-level fine-mapping for isoTWAS and TWAS associations that overlap in a 1 Mb window using methods from the FOCUS framework^67^. We accounted for the correlation between genetically-predicted isoform or gene expression induced by LD and shared prediction weights and control for certain pleiotropic effects. Through Bayesian methods, we estimated the Posterior Inclusion Probability (PIP) for each isoform or gene and defined a credible set of isoforms or genes to explain the signal with 90% confidence.

### Effective sample size

We used the mean χ^2^ statistic from GWAS test results to assess effective sample size. Under the alternative hypothesis, the χ^2^ statistic follows a non-central chi-square distribution with a non-centrality parameter (NCP) that scales with the effective sample size and the squared effect size. As a result, the expected value of the test statistic increases linearly with the effective sample size, making the mean χ^2^ statistic a useful proxy for comparing power across methods or datasets. See **Supplementary Methods** for further details.

### Gene-set enrichment analysis

We conducted gene-set enrichment analysis using enrichR^35^ to investigate the gene ontologies enriched among genes identified by isoTWAS. The analysis queried multiple databases, including GO Biological Process 2023, GO Cellular Component 2023, GO Molecular Function 2023, KEGG 2021 Human, Reactome 2022, and ChEA 2022. ChEA is a curated database of transcription factor targets across multiple ChIP-chip, ChIP-seq, ChIP-PET and DamID assays across a host of tissues and cell-types^82^. We extracted the top 10 most significant terms from each database (FDR-adjusted P < 0.1) and calculated the odds ratios to identify over-represented gene functions.

### Quantitative trait locus mapping and Bayesian colocalization

For the same GTEx tissues that were used in TWAS/isoTWAS, we mapped *cis*-eQTLs for all genes and isoforms using all SNPs within a 1 Mb interval around the transcription start site. We used ordinary least squares to estimate the allelic effect of the SNP (coded as 0, 1, or 2 copies of the risk allele) on gene or isoform expression, adjusted for the same variables used in the original GTEx analyses (five principal components of the genotype matrix, 30-60 PEER factors depending on sample size for the given tissue, age at death, sex). We estimated the eQTL effect sizes, Wald-type standard errors, and P-values.

Next, using GWAS summary statistics for each of the 12 cancer outcomes, gene- and isoform-eQTL summary statistics, and reference LD for European ancestry individuals from the 1000 Genomes Project^76^, we conducted Bayesian colocalization using eCAVIAR^36^ to estimate the CoLocalization Posterior Probability (CLPP) that the same SNP is causal for both cancer risk and the gene or isoform expression. We considered a gene or isoform to colocalize with a GWAS-significant locus if three conditions were met: a SNP has a (1) strong effect on cancer risk (GWAS P < 5 × 10^−8^), (2) strong effect on gene or isoform expression (eQTL P < 10^−6^), and (3) CLPP > 0.01, as is proposed in the original eCAVIAR paper.

### Estimation of expression-mediated SNP heritability

We employed mediated expression score regression (MESC)^9^ to estimate the proportion of total SNP heritability (h^2^) mediated by the *cis*-genetic component of gene or isoform expression levels (h^2^_med_). First, using the genotypes of all SNPs, expression of all genes or isoforms across all 48 tissues in GTEx, and the same covariates used in the eQTL analysis above, we estimated eQTL effect sizes in each tissue using LASSO regression (`./run_mesc.py --compute-expscore-indiv …`). Next, we meta-analyzed expression scores across the 49 tissues and estimated h^2^_med_ for each of the 12 cancer outcomes by using GWAS summary statistics munged to the LD score regression `.sumstats` format. This analysis provided estimates of h^2^, h^2^_med_, and h^2^_med_/h^2^, each with standard errors and P-values for the hypothesis test comparing to the null value of no heritability.

## Supporting information

Supplemental Methods, Results, Figures S1-S37, and Table Legends

## Data Availability

All data produced are available online at https://zenodo.org/records/1104820173 and https://zenodo.org/records/14010391.

https://zenodo.org/records/14010391

## AUTHORS’ CONTRIBUTIONS

YHC: Formal analysis, validation, investigation, visualization, methodology, writing–original draft, writing–review and editing; STB: Formal analysis, writing–review and editing; STH: Formal analysis, writing–review and editing; TH: Formal analysis, writing–review and editing; YY: Formal analysis, validation, visualization, methodology, writing–review and editing; CDH: Methodology, writing–review and editing, funding acquisition; BP: Writing– review and editing, resources, funding acquisition; SL: Writing–review and editing, funding acquisition; AB: Conceptualization, resources, supervision, funding acquisition, investigation, methodology, writing–review and editing

## ACKNOWLEDGEMENTS

Funding was provided by NIH CA293419 and CA194393. The funder had no role in study design, data collection, analysis, or interpretation, or writing of the article.

