## Supplemental Methods, Results, Figures S1-S37, and Table Legends for "Isoform-level analyses of 6 cancers uncover extensive genetic risk mechanisms undetected at the gene-level"

**SUPPLEMENTAL MATERIALS**

**Table of Contents**

| Supplemental Methods |  | 1 |
| --- | --- | --- |
| Supplemental Results |  | 1 |
| Supplemental Figures |  | 4 |
| Supplemental Table, Data Legends, and Reference |  | 41 |

**SUPPLEMENTAL METHODS**

**Overview of isoTWAS**

We used univariate and multivariate penalized regressions (**Supplemental Figure 1A**) to train gene and isoform expression models and map gene- and isoform-level risk associations for the 12 cancer outcomes using a weighted burden test (**Supplemental Figure 1B**)^1,2^. In isoTWAS, we inferred gene-level associations by combining isoform associations via aggregated Cauchy association^3^. False discovery rate is controlled for via a Benjamini-Hochberg procedure, and family-wise error rates are controlled via Shaffer’s modified sequentially rejective Bonferroni (MSRB) for correlated isoforms. Post-hoc analyses include permutation tests and fine-mapping to identify isoform sets driving significant associations and credible sets of genes or isoforms (**Supplemental Figure 1C**).

**Effective Sample Size**

In GWAS, when a variant has a non-zero effect, the association test statistic $Z$ will follow a normal distribution $Z \sim N(\frac{\beta}{SE},1)$ under the alternative hypothesis. Squaring $Z$ yields $Z^{2} \sim\chi_{1}^{2}(\lambda)$where the test statistic follows a non-central chi-square distribution with 1 degree of freedom and non-centrality parameter (NCP) $\lambda= \left( \frac{\beta^{2}}{{SE}^{2}} \right)$. This non-centrality parameter λ is directly proportional to the effective sample size. In typical association tests, the standard error $SE(\hat{\beta})$decreases with increasing sample size $n_{eff}$, such that λ∝$n_{eff}\cdot\beta^{2}$. As a result, the mean of the $\chi^{2}$ statistic, ${E(\chi}^{2})$ $=1+\lambda$, increases linearly with effective sample size. Therefore, increases in the observed mean $\chi^{2}$ statistic can be interpreted as increases in statistical power or effective sample size.

**Rare variant analysis in UK Biobank**

We used whole exome sequencing data from the UK Biobank^4^ (N_cases_ = 8,023 and N_cancer-free-controls_ = 39,121) and conducted a rare variant analysis (MAF < 0.5%) on *BABAM1* for BRCA. After obtaining the sequence data of all UK Biobank samples (200K release), we conducted joint genotype calling using DeepVariant and GLnexus, followed by sample and variant quality control using XPAT^5^. Using the Variant Annotation, Analysis, and Search Tool (VAAST2)^6,7^, a probabilistic tool that assigns a pathogenicity weight to each rare variant, we compiled a burden test for each transcript-isoform and exon of the gene. Here, we considered nominal P < 0.05 as a suggestive association indicating increased pathogenic rare variant signal from a transcriptomic feature.

**SUPPLEMENTAL RESULTS**

**isoTWAS associations with isoform-eQTL colocalizations**

We summarize 6 additional loci that show a shared pattern: strong GWAS signal within and/or near the gene body, no gene-eQTL signal, and a colocalized (CLPP > 0.01) isoform-eQTL signal for an isoform prioritized by isoTWAS and post-hoc isoform-level fine-mapping.

1. We find 6 isoforms of *FDPS* (Chromosome 1q22, s_het_ = 0.007, 21 total isoforms) associated with BRCA, using models trained in lymphocytes. Fine-mapping prioritized ENST00000492244.5 in the 90% credible set with PIP = 1.0 and CLPP = 0.12. There are shared exons across many of these isoforms, especially with ENST00000471117.5 (CLPP=0.04) and ENST00000489324.1 (CLPP=0.07). The lead isoform-eQTL rs11264361 has a significant protective effect on BRCA risk and strong positive associations with ENST00000492244.5, ENST00000489324.1, and ENST00000471117.5 expression (**Supplemental Figure S24**).
2. We find 1 isoform of *L3MBTL3* (Chromosome 6q23.1, s_het_ = 0.10, 10 total isoforms) associated with ER- BRCA, using models trained in fibroblasts. Fine-mapping prioritized ENST00000531313.1 in the 90% credible set with PIP = 1.0 and CLPP = 0.03. The lead isoform-eQTL rs6569648 has a significant positive effect on ER- BRCA risk and a strong positive association with ENST00000531313.1 expression (**Supplemental Figure S25**).
3. We find 2 isoforms of *CASP8* (Chromosome 2q33.1, s_het_ = 0.017, 25 total isoforms) associated with ER- BRCA, using models trained in ovary. Fine-mapping prioritized ENST00000392266.7 in the 90% credible set with PIP = 1.0 and CLPP = 0.33. There are shared exons between these 2 isoforms. However, the lead isoform-eQTL rs10931936 has a significant positive effect on ER- BRCA risk and strong positive associations with ENST00000392266.7, yet no effect on ENST00000264275.9 expression (**Supplemental Figure S26**).
4. We find 11 isoforms of *TMBIM1* (Chromosome 2q35, s_het_ = 0.006, 23 total isoforms) associated with BRCA, using models trained in subcutaneous adipose tissue. Fine-mapping prioritized ENST00000437694.6 in the 90% credible set with PIP = 1.0 and CLPP = 0.05. The complexity of the exon structure across these isoforms is reflected in the effects of the lead isoform-QTL rs2382817 on isoform expression with large effects on multiple isoforms, including ENST00000437694.6 (**Supplemental Figure S27**).
5. We find 4 isoforms of *GGCX* (Chromosome 2p11.2, s_het_ = 0.011, 25 total isoforms) associated with PRCA, using models trained in subcutaneous adipose tissue. Fine-mapping prioritized ENST00000430215.7 (PIP = 0.21, CLPP = 0.69) and ENST00000473665.1 (PIP = 0.19, CLPP = 0.15) in the 90% credible set. There are shared exons across these 2 isoforms, with additional shared exons across the 2 isoforms that were not prioritized in fine-mapping. The lead isoform-eQTL rs6757263 has a significant positive effect on PRCA risk and strong positive associations with both ENST00000430215.7 and ENST00000473665.1 (**Supplemental Figure S28**).

We find 3 isoforms of *SRP14* (Chromosome 15q22, s_het_ = 0.19, 8 total isoforms) associated with UCEC, using models trained in vaginal tissue. Fine-mapping did not prioritize any of these isoforms, however we find CLPP > 0.01 across all three isoforms. Most exons are shared across these isoforms, with isoform-eQTL effects largely in the same direction for all three isoforms, mimicking the protective effects on UCEC risk (**Supplemental Figure S29**).

In total, these results underscored the increased resolution that can be achieved to contextualize cancer risk GWAS loci without additional sequencing costs. However, these results also highlight a methodological opportunity in leveraging shared exon structure to further resolve credible sets of causal isoforms underlying GWAS loci.

**Rare Variants Analysis on *BABAM1***

We specifically focused on *BABAM1* due to its distinct transcript structure and the sufficient statistical power available in the UK Biobank whole exome sequencing data. To further investigate rare variant effects on BRCA, we used whole exome sequencing data from the UK Biobank and conducted a rare variant analysis (MAF < 0.5%) on *BABAM1*. Using VAAST2, we identify three significant BRCA-associated isoforms for *BABAM1* (P < 0.05; **Supplemental Tables S11**), indicating that relevant risk variants in these isoforms are enriched in cases compared to controls. Additionally, exon-specific results show that significant rare variants are predominantly located at the 5' end of the transcript and within the first exon at the 3' end (**Supplemental Figure 36; Supplemental Tables S11**).

**SUPPLEMENTAL FIGURES**
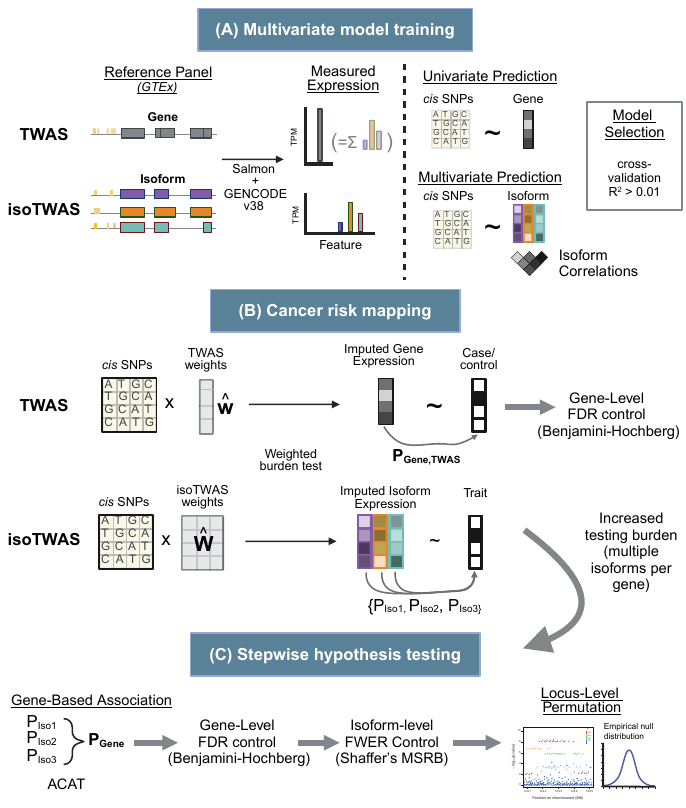

**Figure S1**: *Overview of isoform- and gene-level TWAS*. **(A)** Using multi-tissue GTEx data, isoform- and gene-level expression was quantified using Salmon and GENCODE v38 annotations. Univariate and multivariate predictive models for gene and isoform expression were trained. **(B)** Using the weighted burden test, predictive models of gene and isoform expression were used to map gene- and isoform-level cancer risk associations. For gene-level TWAS associations, FDR is controlled to 5% via Benjamini-Hochberg. **(C)** As isoTWAS presents an increased testing burden, stepwise hypothesis testing is conducted. Isoform P-values are aggregated to the gene-level via aggregated Cauchy aggregation. FDR is controlled at 5% via Benjamini-Hochberg procedure on gene-level P-values. For significantly-associated genes, FWER is controlled to 5% for correlated isoform associations using Shaffer’s MSRB procedure. Locus-level permutation is conducted to control of local LD patterns. Modified from Bhattacharya et al 2023.

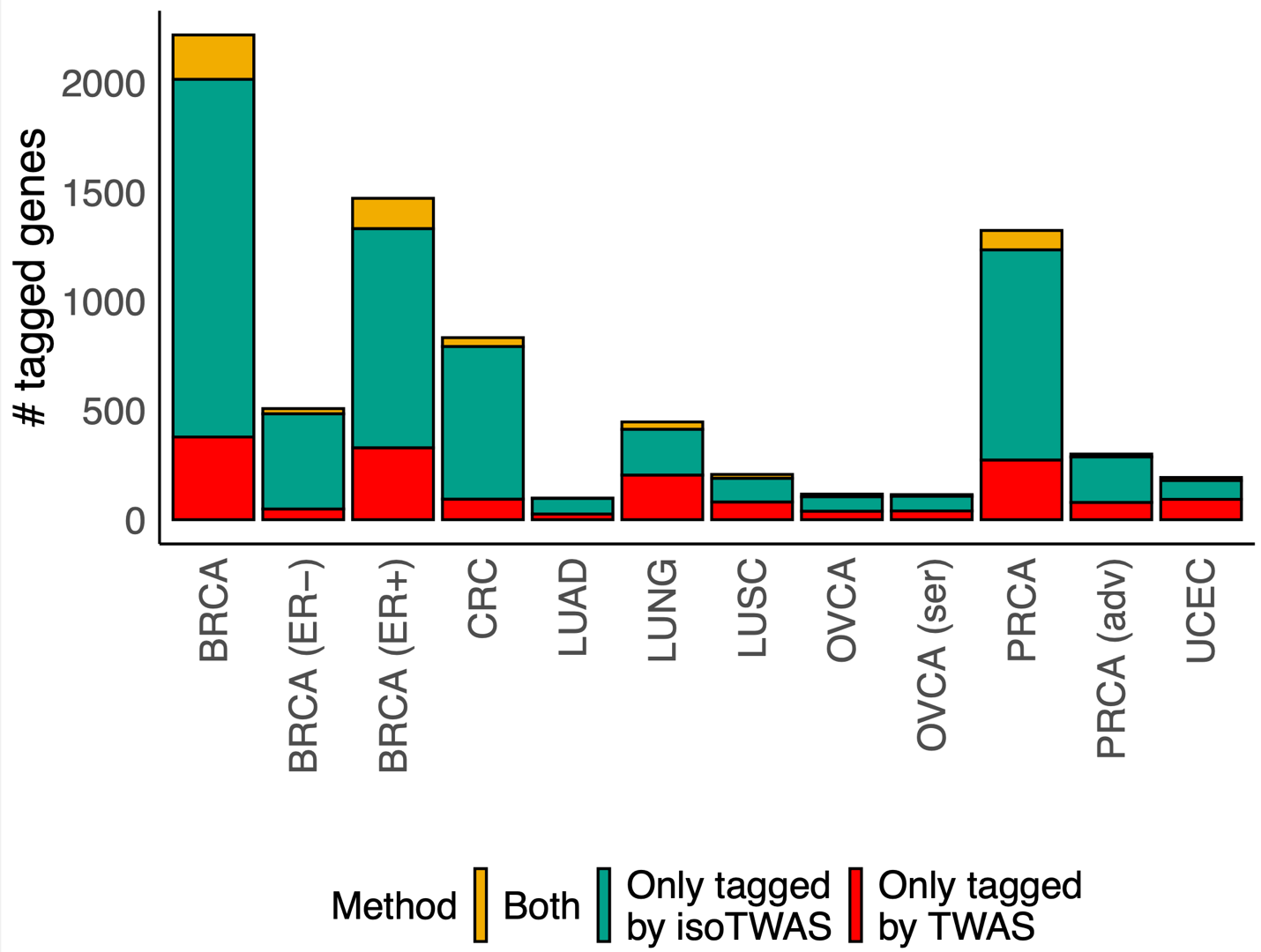

**Figure S2**: *The number of unique genes that tagged by isoTWAS (green), TWAS (red), or both in common (gold).*

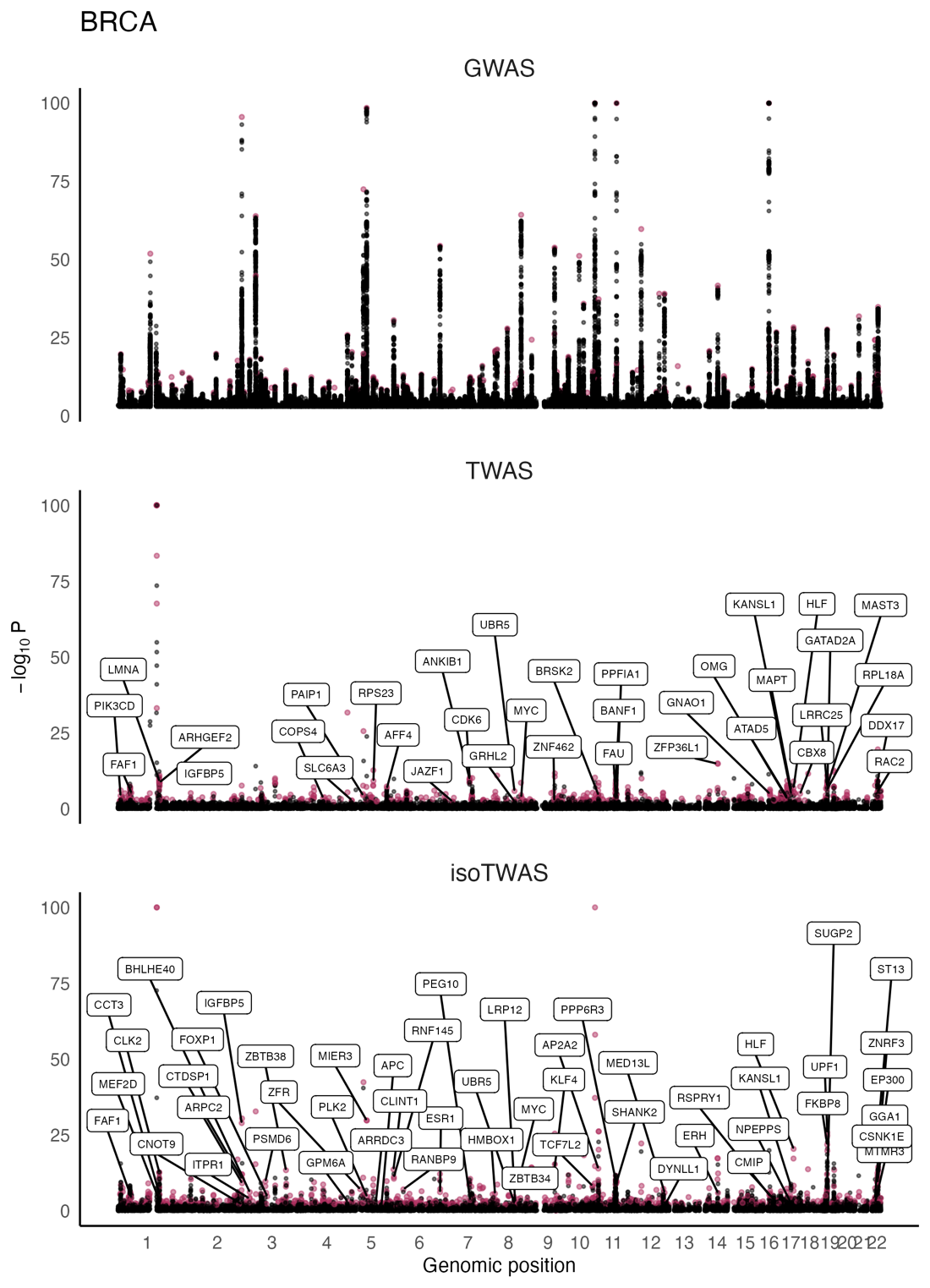

**Figure S3**: Manhattan plots for BRCA GWAS (top) and gene associations from TWAS (middle), and isoTWAS (bottom). Points are colored if the gene is transcriptome-wide significant in TWAS or isoTWAS and labelled if it is within 1 Mb of a GWAS-significant SNP and has s_het_ > 0.10.

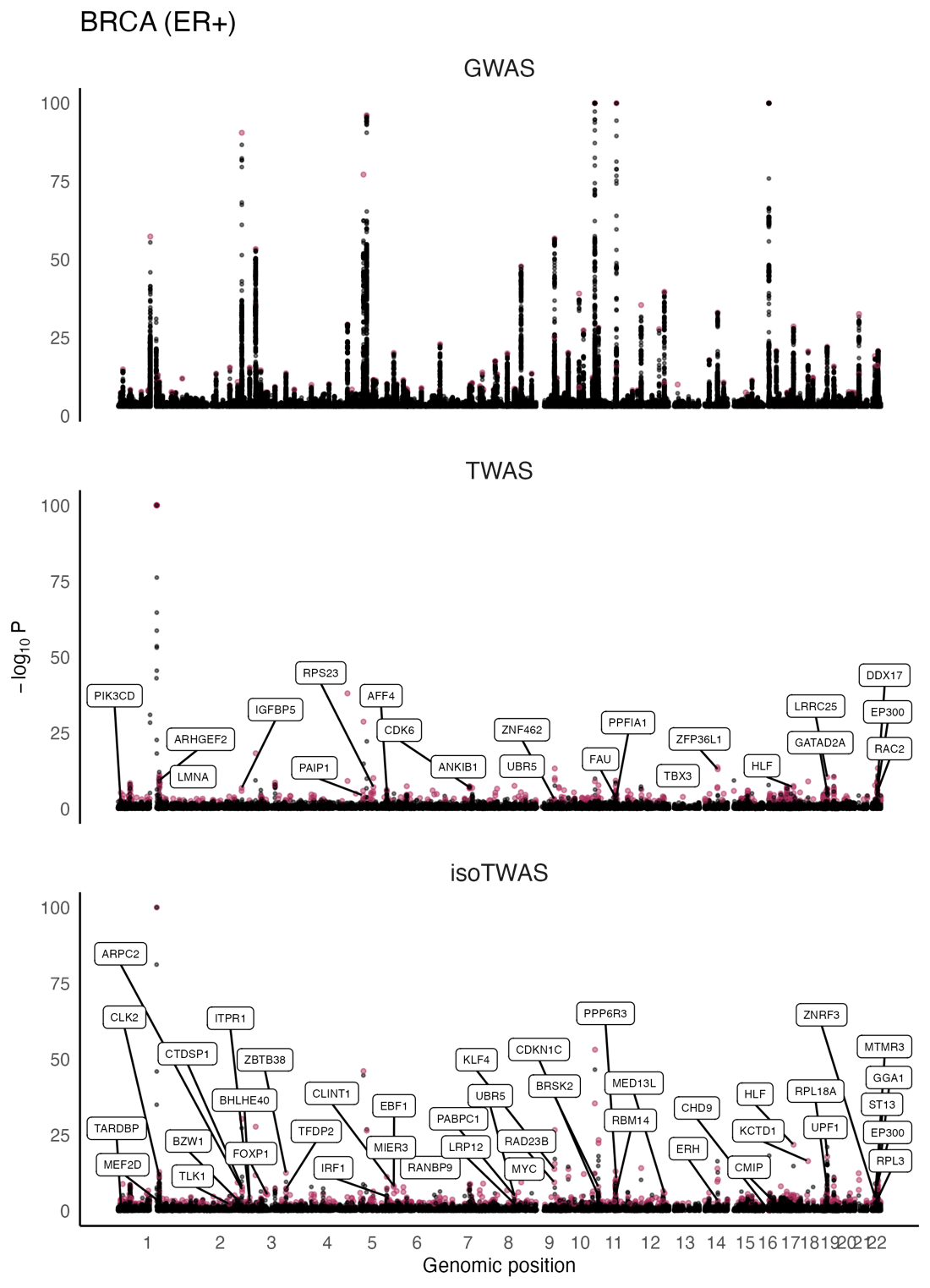
**Figure S4**: Manhattan plots for ER+ BRCA GWAS (top) and gene associations from TWAS (middle), and isoTWAS (bottom). Points are colored if the gene is transcriptome-wide significant in TWAS or isoTWAS and labelled if it is within 1 Mb of a GWAS-significant SNP and has s_het_ > 0.10.

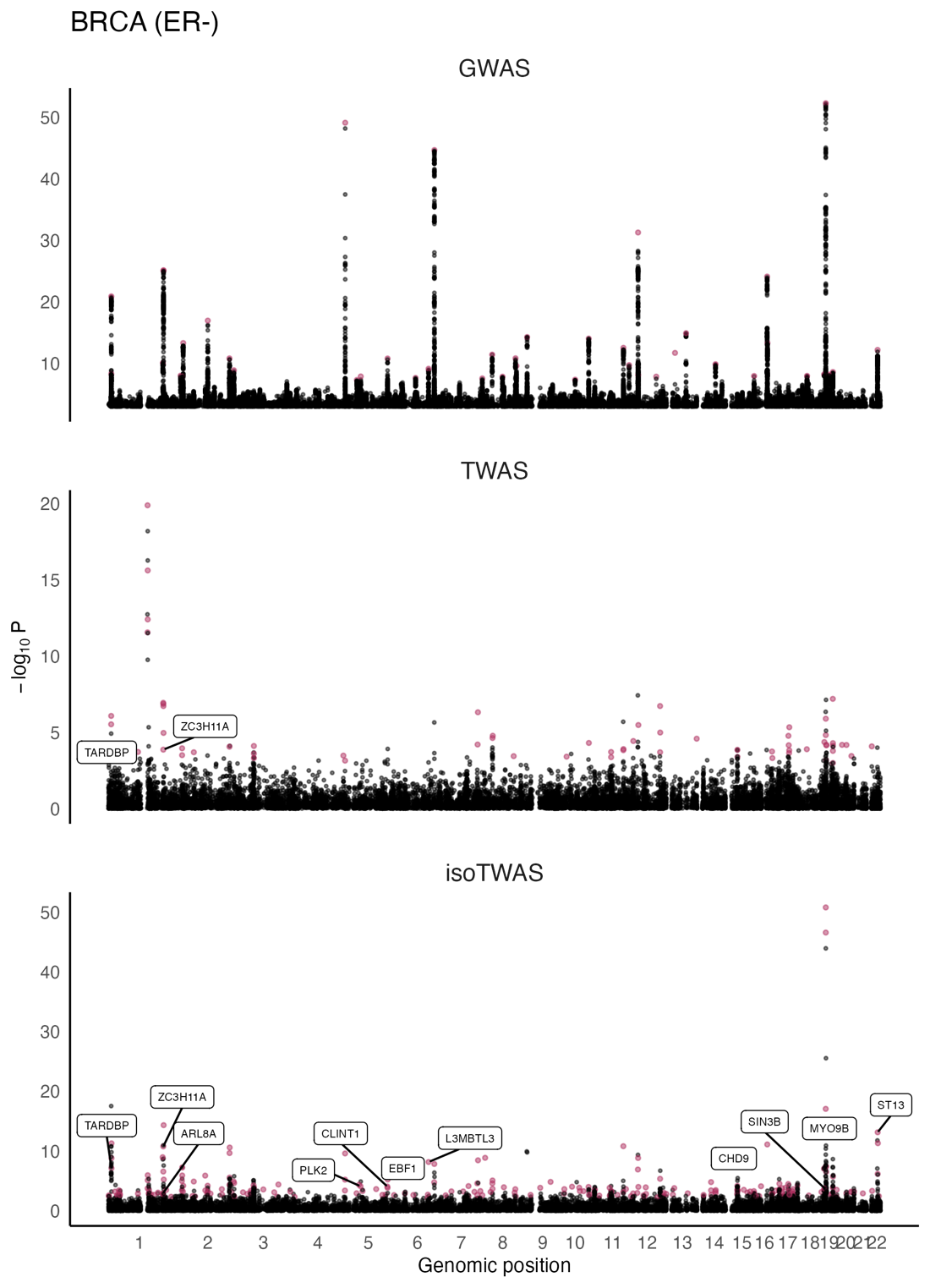
**Figure S5**: Manhattan plots for ER- BRCA GWAS (top) and gene associations from TWAS (middle), and isoTWAS (bottom). Points are colored if the gene is transcriptome-wide significant in TWAS or isoTWAS and labelled if it is within 1 Mb of a GWAS-significant SNP and has s_het_ > 0.10.

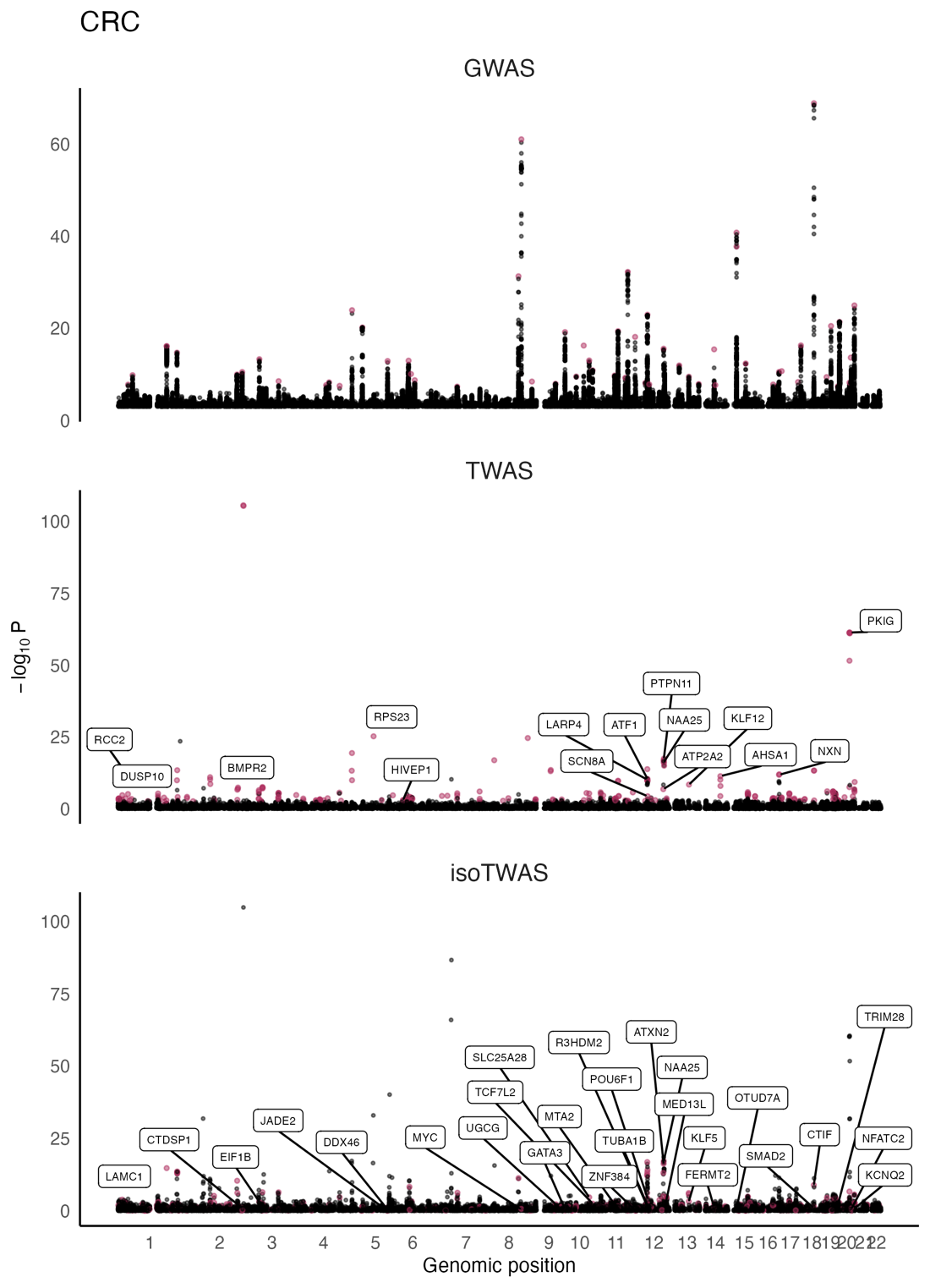
**Figure S6**: Manhattan plots for CRC GWAS (top) and gene associations from TWAS (middle), and isoTWAS (bottom). Points are colored if the gene is transcriptome-wide significant in TWAS or isoTWAS and labelled if it is within 1 Mb of a GWAS-significant SNP and has s_het_ > 0.10.

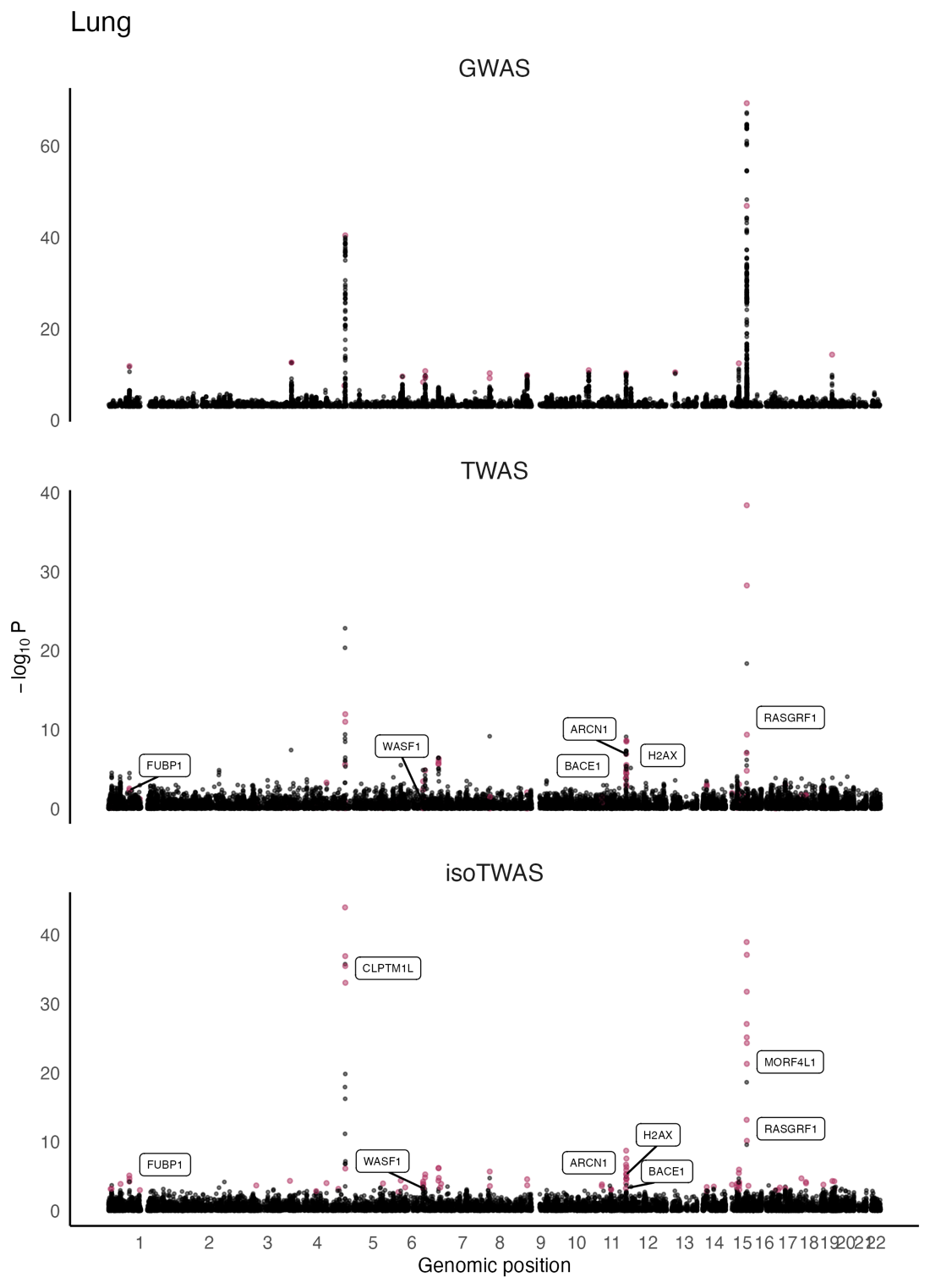
**Figure S7**: Manhattan plots for LUNG GWAS (top) and gene associations from TWAS (middle), and isoTWAS (bottom). Points are colored if the gene is transcriptome-wide significant in TWAS or isoTWAS and labelled if it is within 1 Mb of a GWAS-significant SNP and has s_het_ > 0.10.

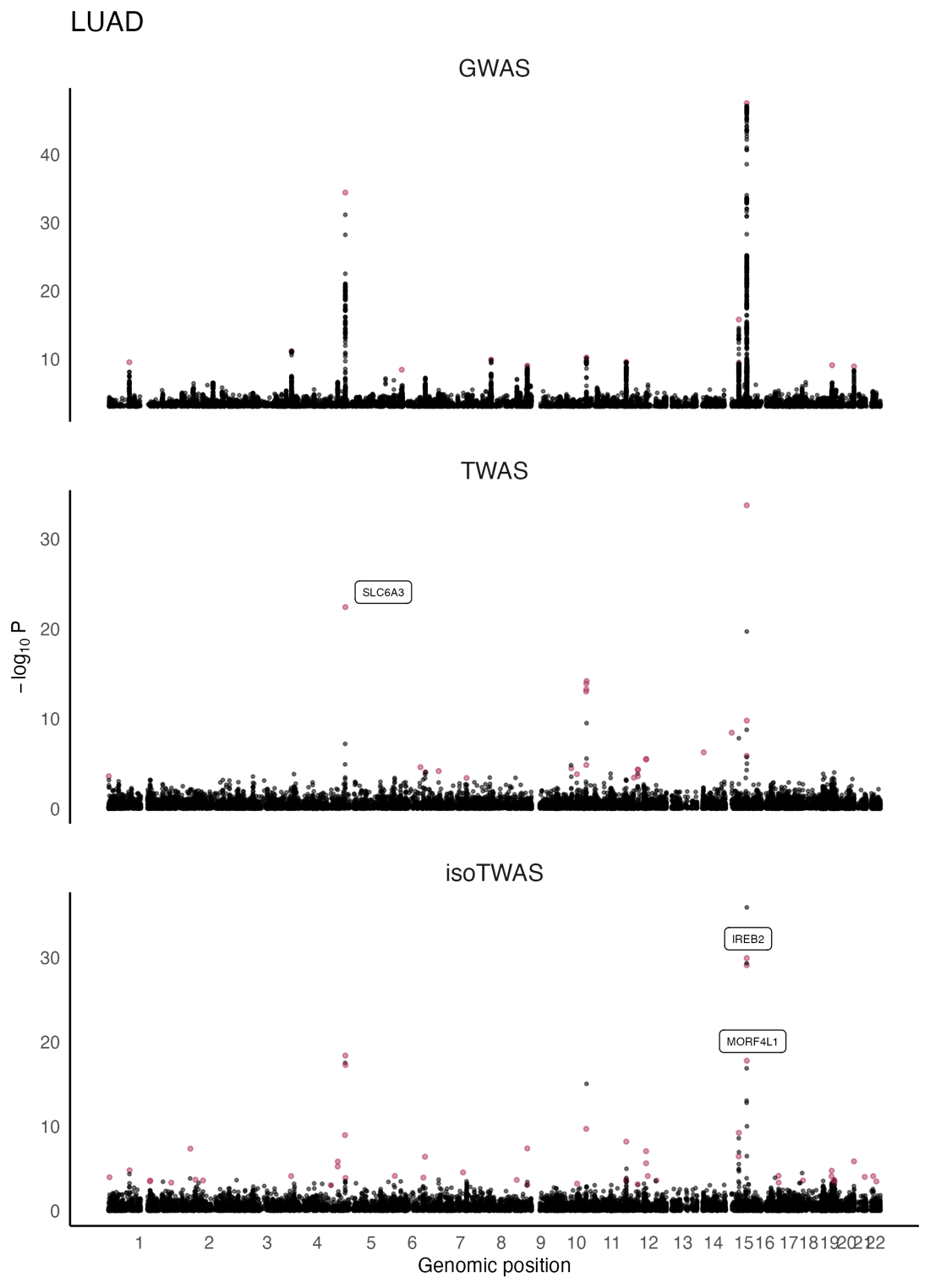
**Figure S8**: Manhattan plots for LUAD GWAS (top) and gene associations from TWAS (middle), and isoTWAS (bottom). Points are colored if the gene is transcriptome-wide significant in TWAS or isoTWAS and labelled if it is within 1 Mb of a GWAS-significant SNP and has s_het_ > 0.10.

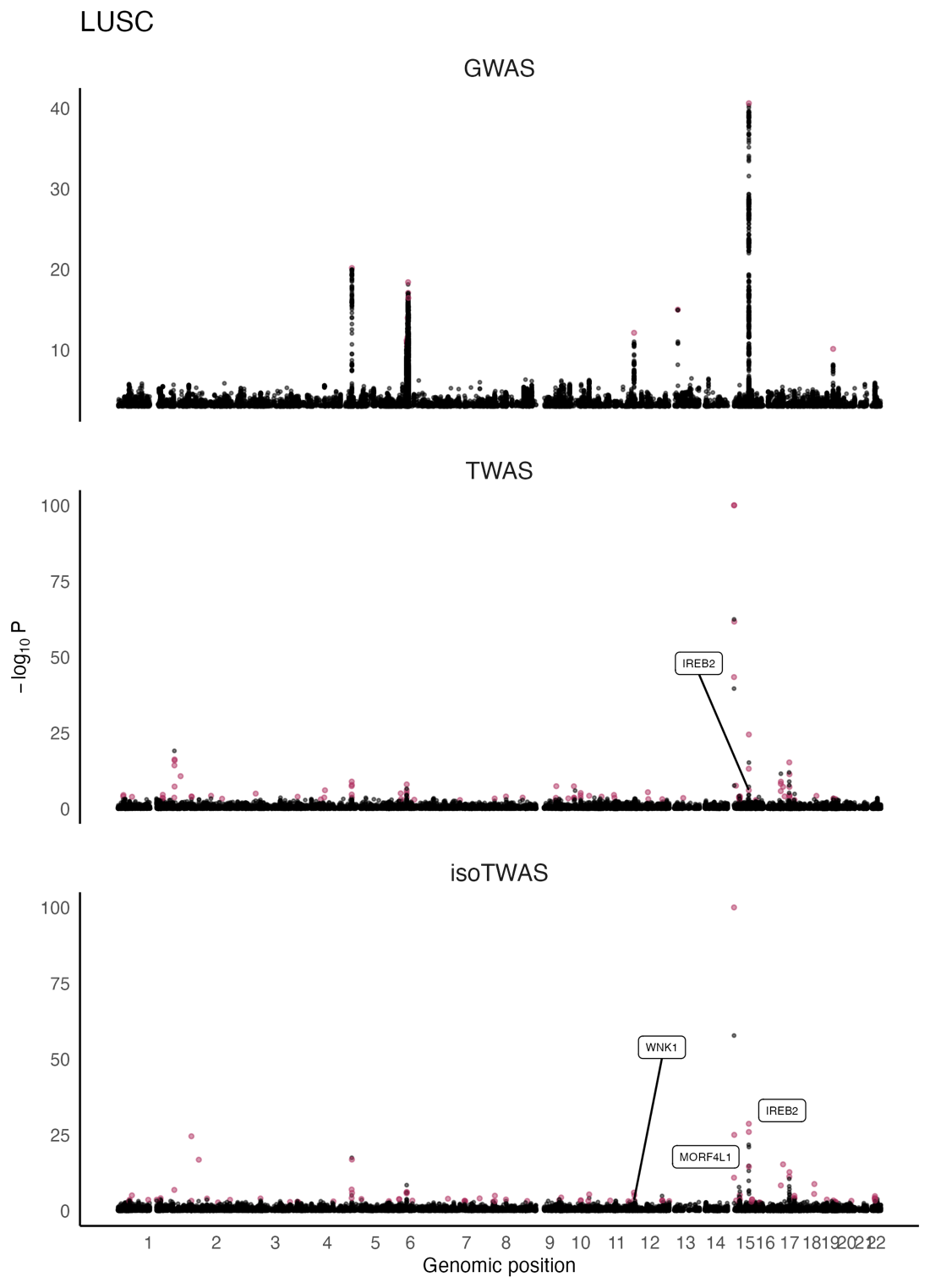
**Figure S9**: Manhattan plots for LUSC GWAS (top) and gene associations from TWAS (middle), and isoTWAS (bottom). Points are colored if the gene is transcriptome-wide significant in TWAS or isoTWAS and labelled if it is within 1 Mb of a GWAS-significant SNP and has s_het_ > 0.10.

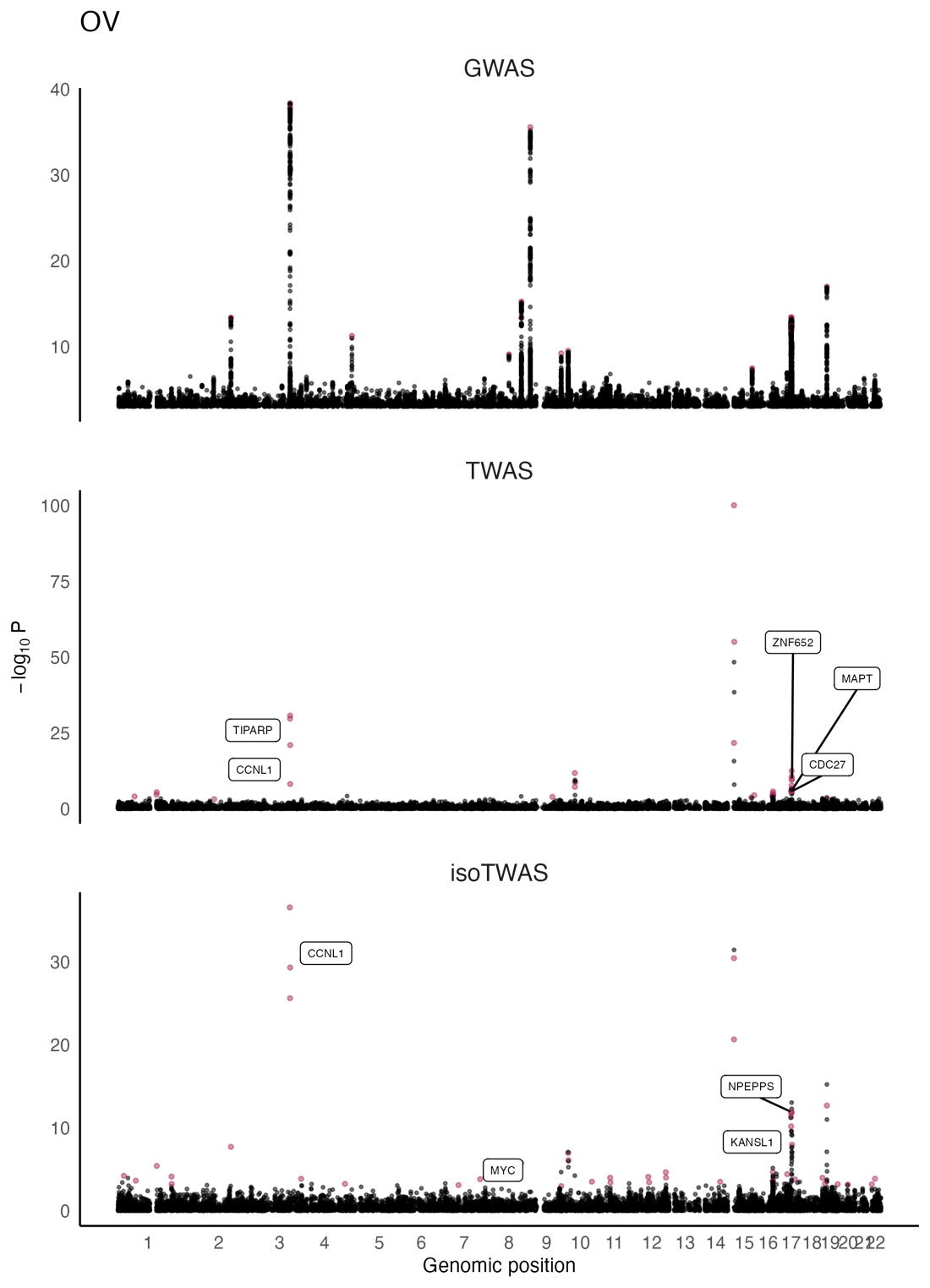
**Figure S10**: Manhattan plots for OVCA GWAS (top) and gene associations from TWAS (middle), and isoTWAS (bottom). Points are colored if the gene is transcriptome-wide significant in TWAS or isoTWAS and labelled if it is within 1 Mb of a GWAS-significant SNP and has s_het_ > 0.10.

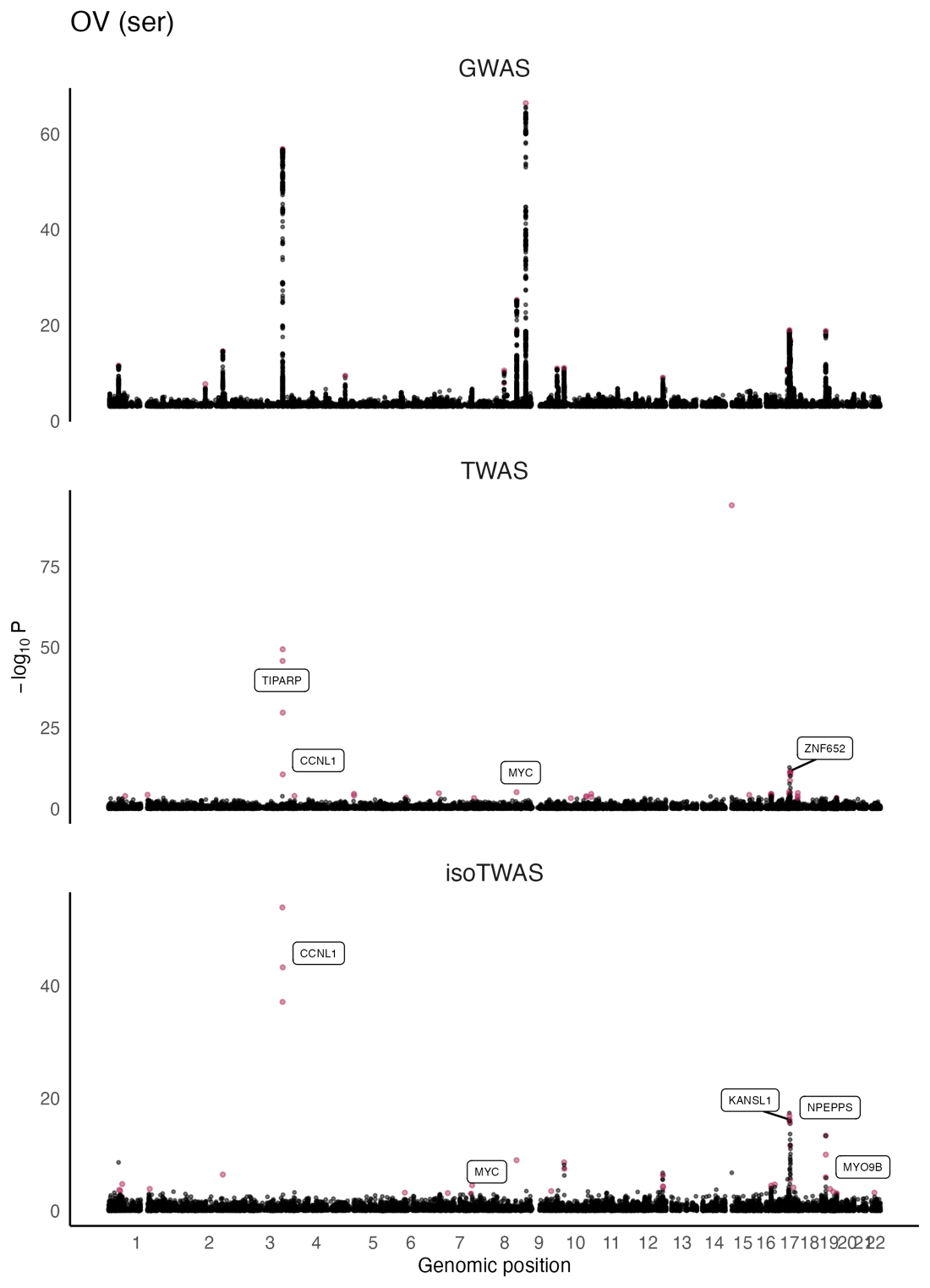
**Figure S11**: Manhattan plots for OVCA ser GWAS (top) and gene associations from TWAS (middle), and isoTWAS (bottom). Points are colored if the gene is transcriptome-wide significant in TWAS or isoTWAS and labelled if it is within 1 Mb of a GWAS-significant SNP and has s_het_ > 0.10.

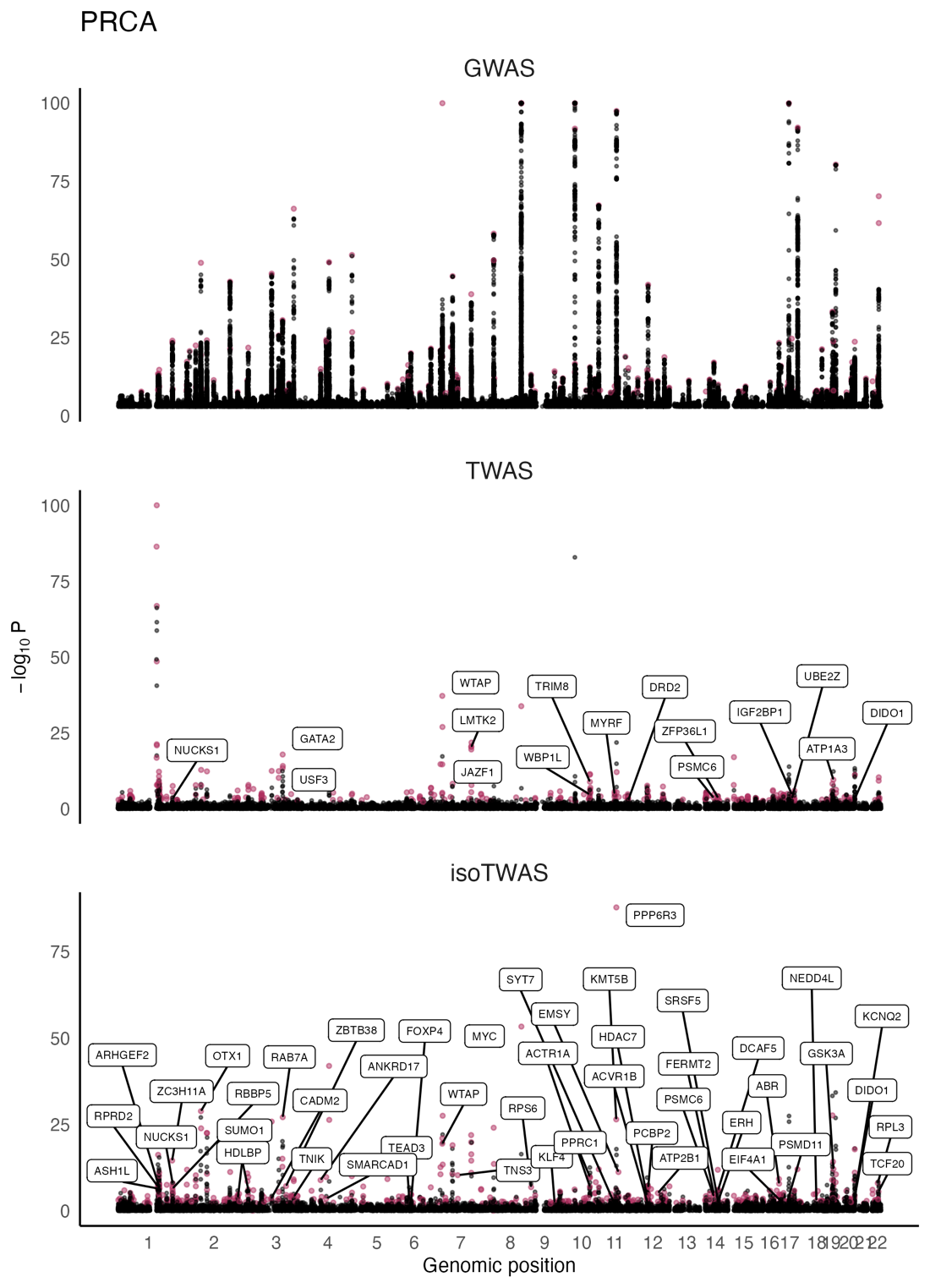
**Figure S12**: Manhattan plots for PRCA GWAS (top) and gene associations from TWAS (middle), and isoTWAS (bottom). Points are colored if the gene is transcriptome-wide significant in TWAS or isoTWAS and labelled if it is within 1 Mb of a GWAS-significant SNP and has s_het_ > 0.10.

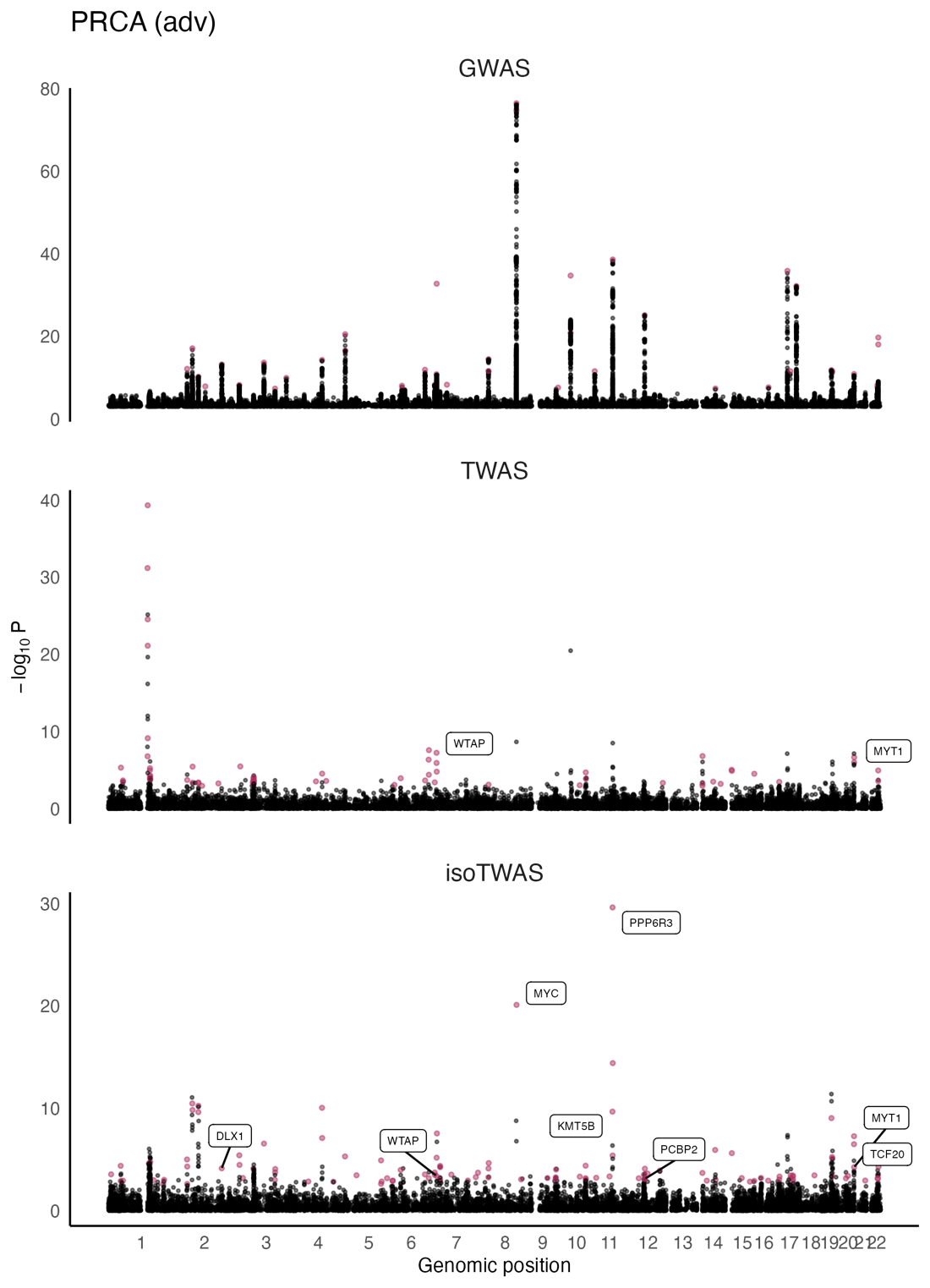
**Figure S13**: Manhattan plots for Adv PRCA GWAS (top) and gene associations from TWAS (middle), and isoTWAS (bottom). Points are colored if the gene is transcriptome-wide significant in TWAS or isoTWAS and labelled if it is within 1 Mb of a GWAS-significant SNP and has s_het_ > 0.10.

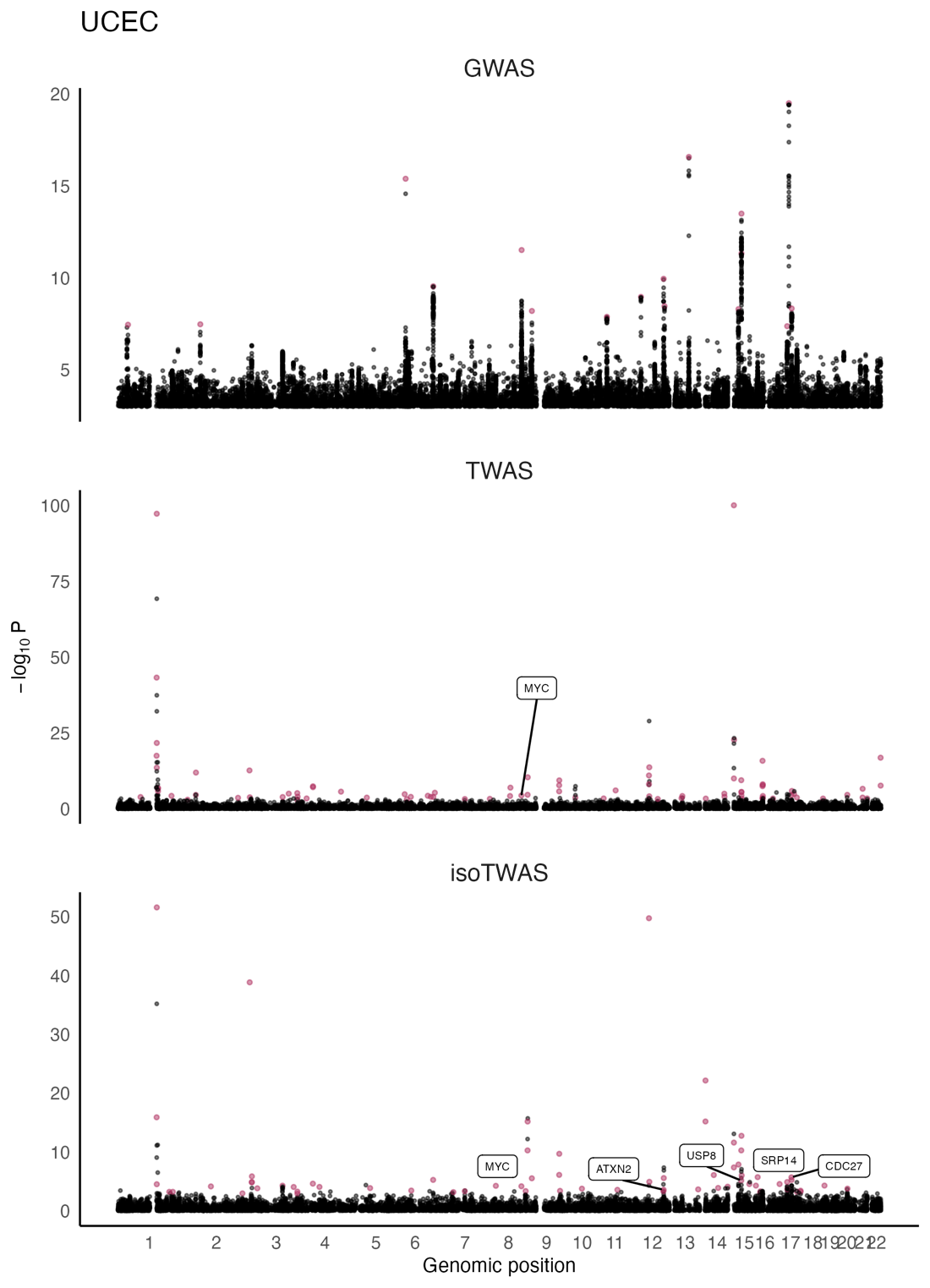
**Figure S14**: Manhattan plots for UCEC GWAS (top) and gene associations from TWAS (middle), and isoTWAS (bottom). Points are colored if the gene is transcriptome-wide significant in TWAS or isoTWAS and labelled if it is within 1 Mb of a GWAS-significant SNP and has s_het_ > 0.10.

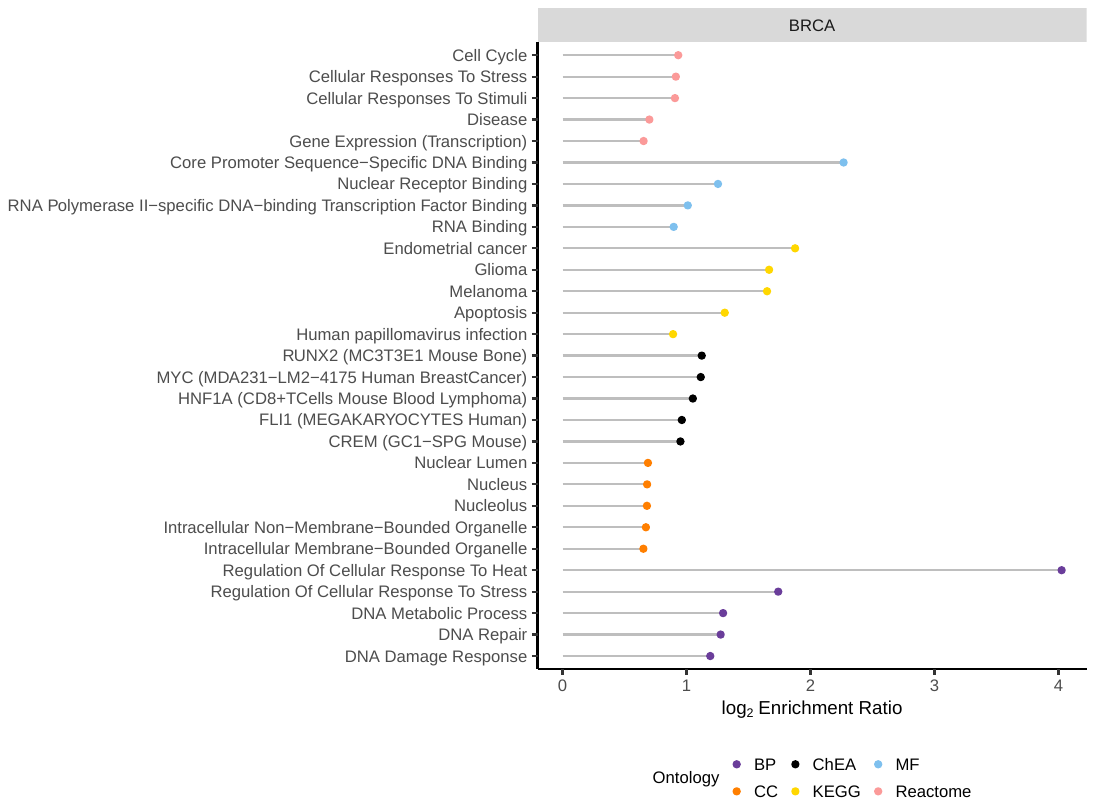
**Figure S15**: Log enrichment ratio (X-axis) of enriched pathways (Y-axis) for isoTWAS-prioritized genes associated with BRCA (including ER+ and ER-), colored by ontology.

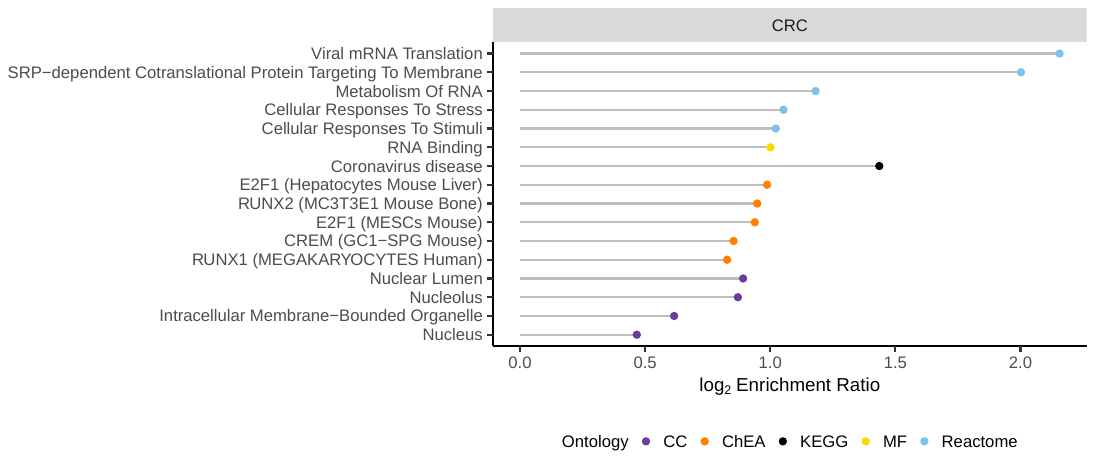
**Figure S16**: Log enrichment ratio (X-axis) of enriched pathways (Y-axis) for isoTWAS-prioritized genes associated with CRC, colored by ontology.

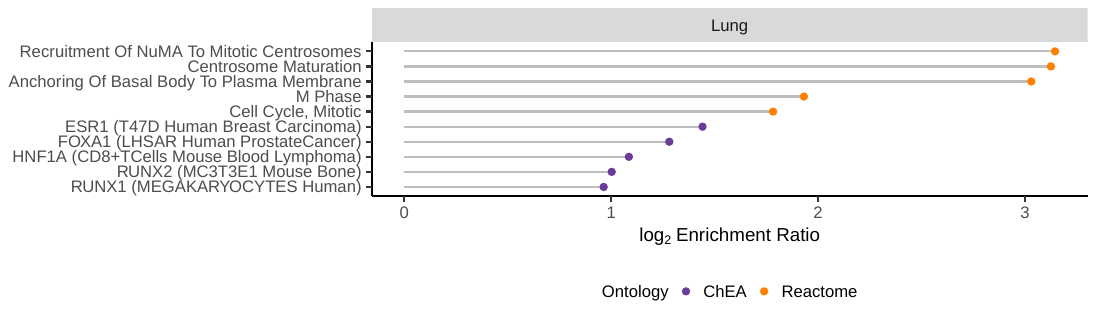
**Figure S17**: Log enrichment ratio (X-axis) of enriched pathways (Y-axis) for isoTWAS-prioritized genes associated with lung cancer (including LUAD and LUSC), colored by ontology.

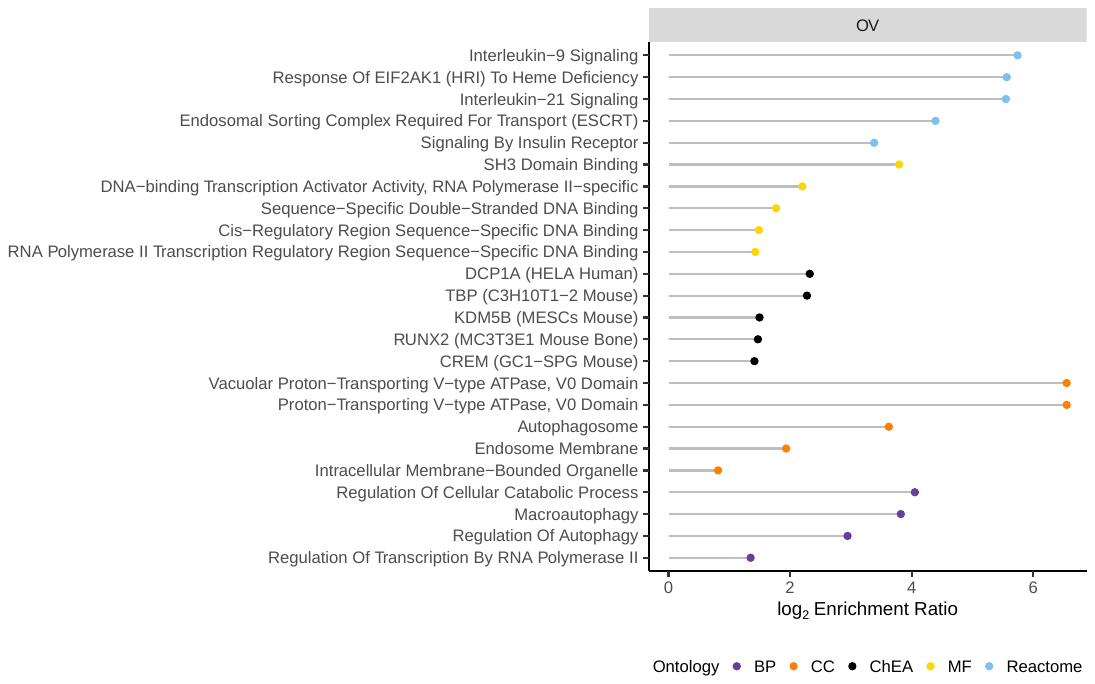
**Figure S18**: Log enrichment ratio (X-axis) of enriched pathways (Y-axis) for isoTWAS-prioritized genes associated with OVCA (including serous subtype), colored by ontology.

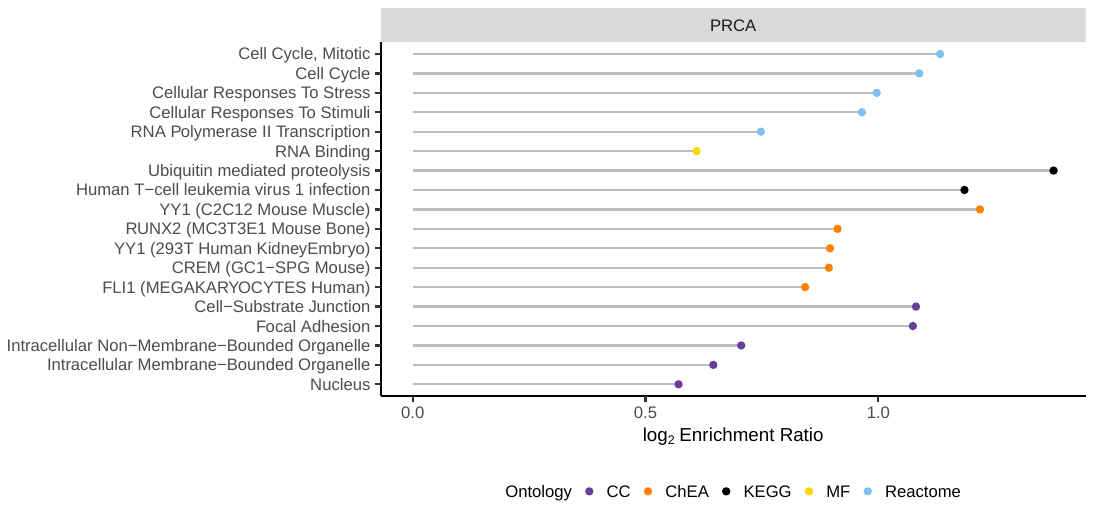
**Figure S19**: Log enrichment ratio (X-axis) of enriched pathways (Y-axis) for isoTWAS-prioritized genes associated with PRCA (including advanced subtype), colored by ontology.

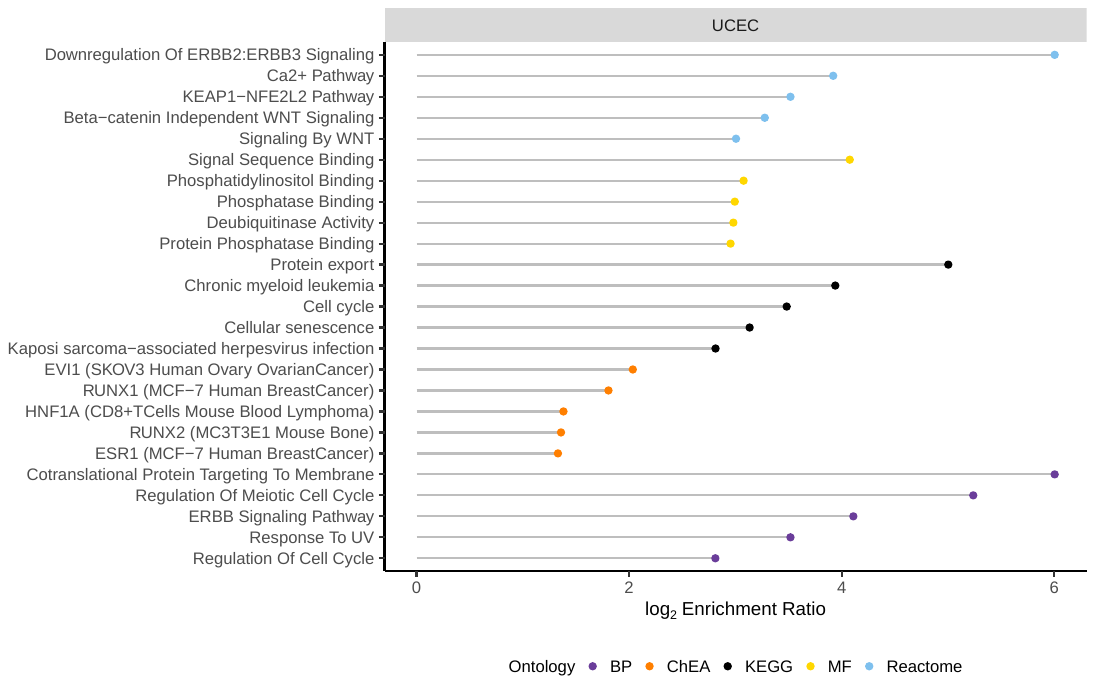
**Figure S20**: Log enrichment ratio (X-axis) of enriched pathways (Y-axis) for isoTWAS-prioritized genes associated with UCEC, colored by ontology.

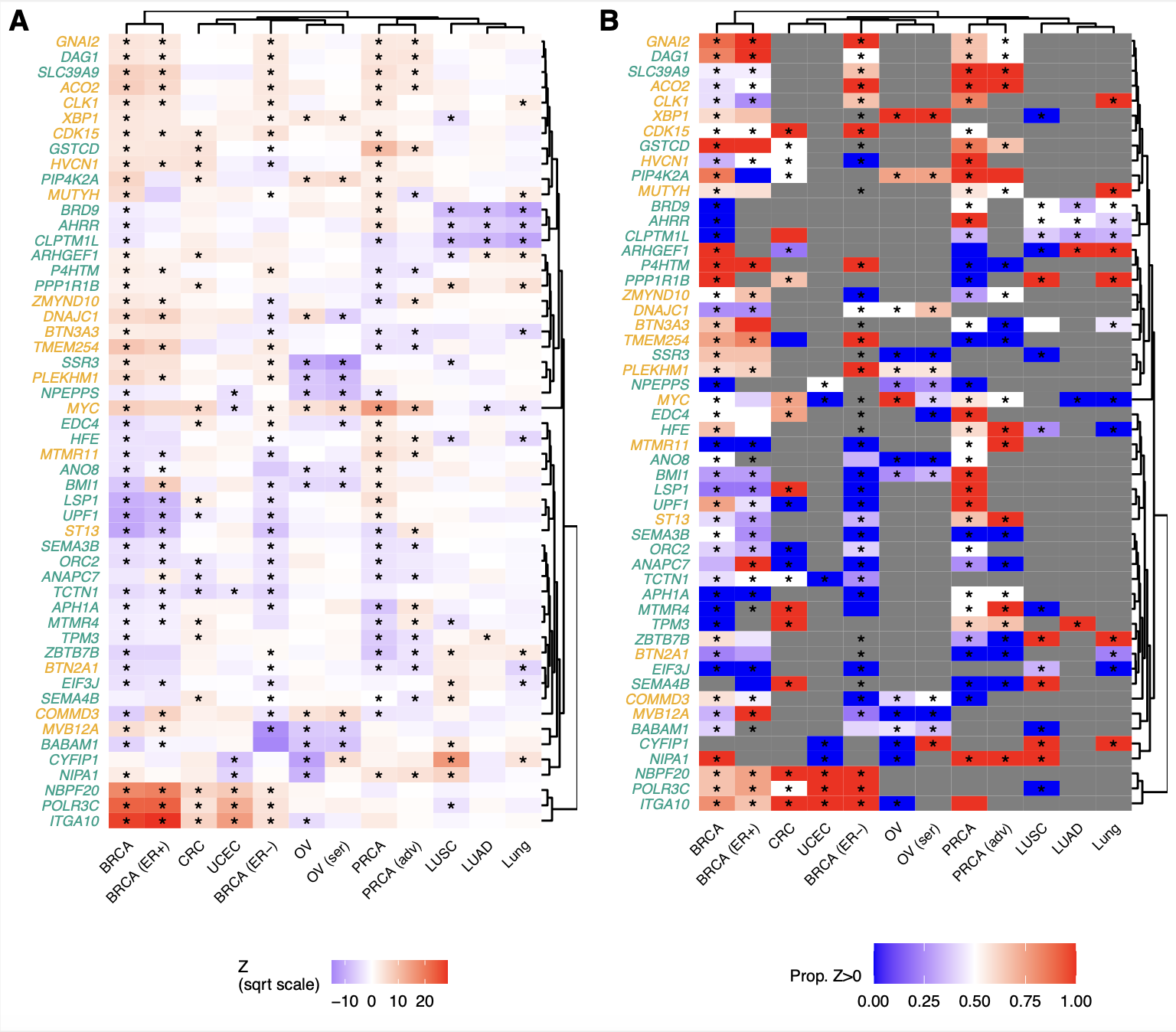

**Figure S21**: (A) Scaled gene-level isoTWAS Z-scores for 52 genes with isoform-level risk associations across at least 5 cancer outcomes out of 12 total. (B) Proportion of transcripts with positive direction of effect for each gene shown in (A).

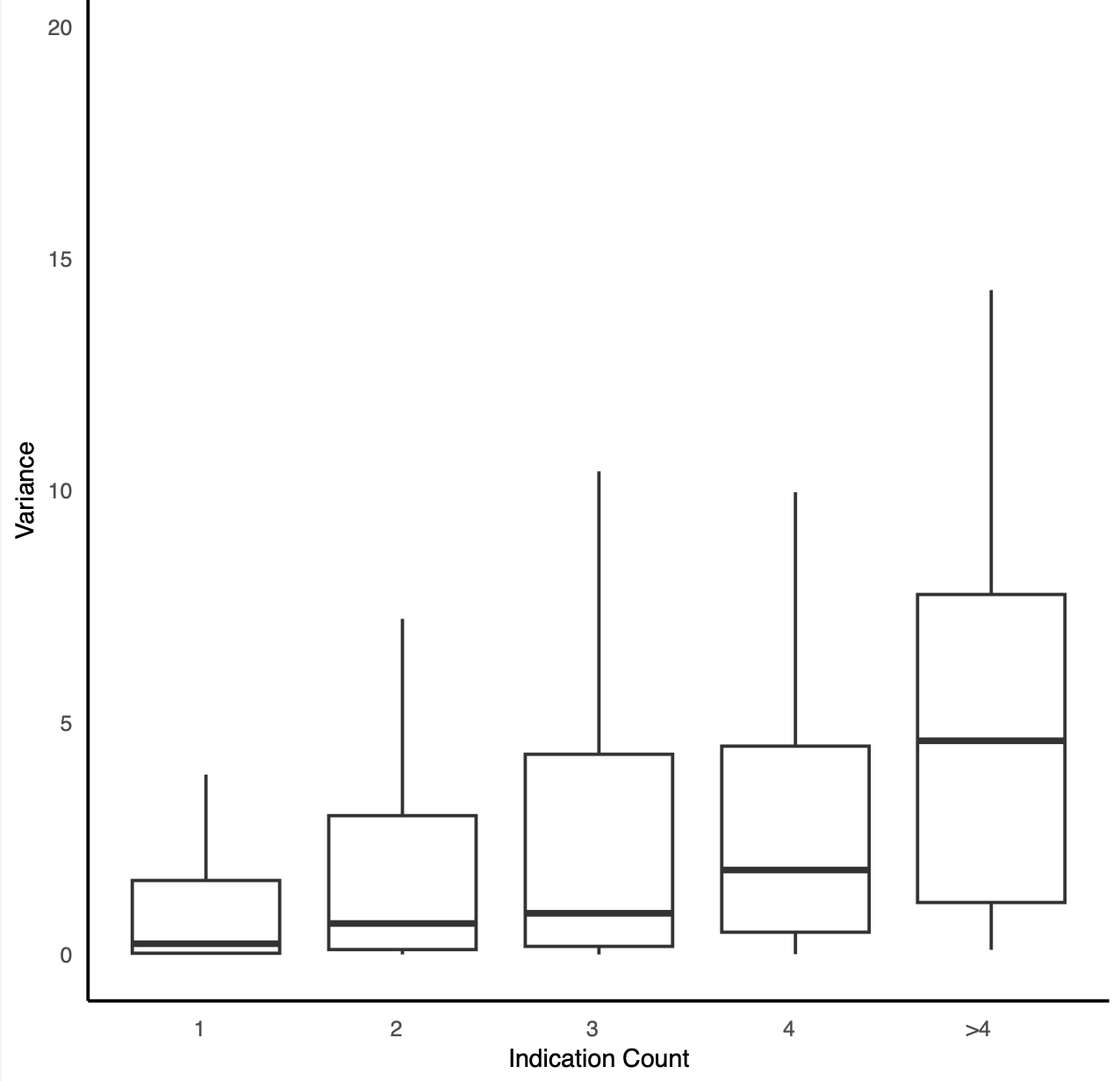

**Figure S22**: *Genes linked to more cancers tend to have higher expression variability across tissues.* The x-axis represents the number of cancers each gene is associated with, and the y-axis shows the corresponding expression variance across tissues. ANOVA was used to estimate variance attributable to tissue, while accounting for between-transcript variability.

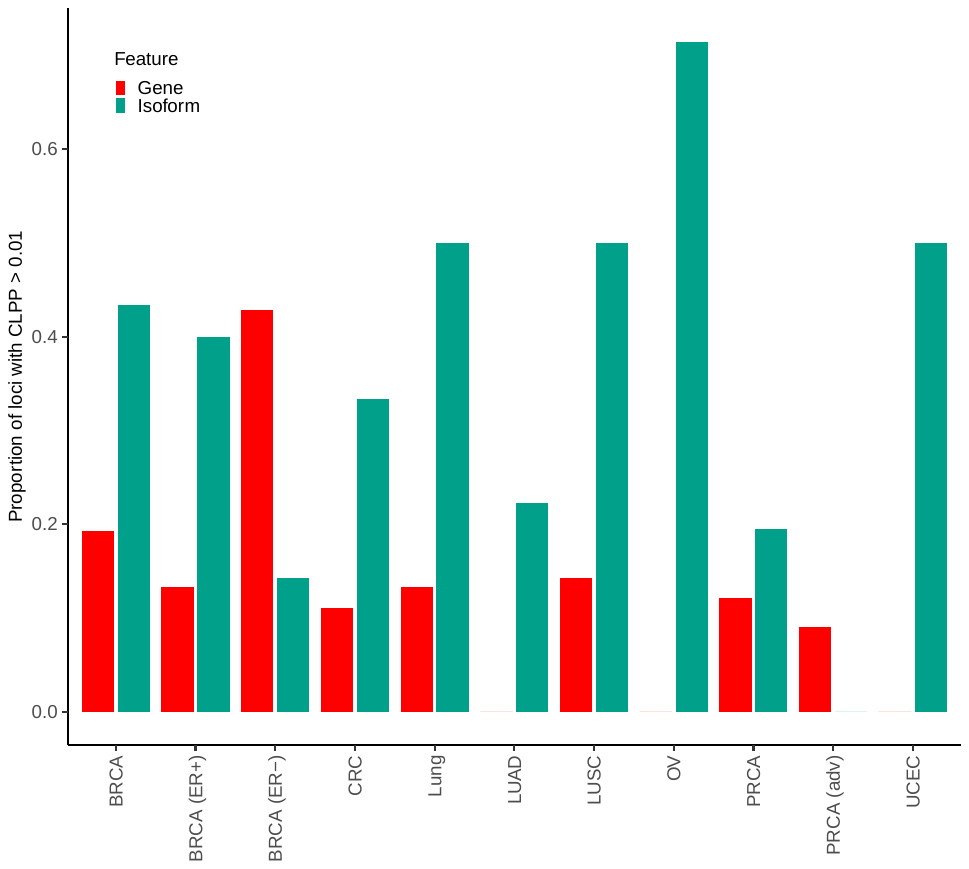

**Figure S23**: *Isoform expression colocalizes far better with cancer risk loci from GWAS than gene expression.* Across 12 cancer indications (X-axis), the proportion of GWAS loci with a SNP P < 5x10^-8^ that colocalizes with CLPP > 0.01 with a gene- (red) or isoform-eQTL (green) with P < 10^-6^.

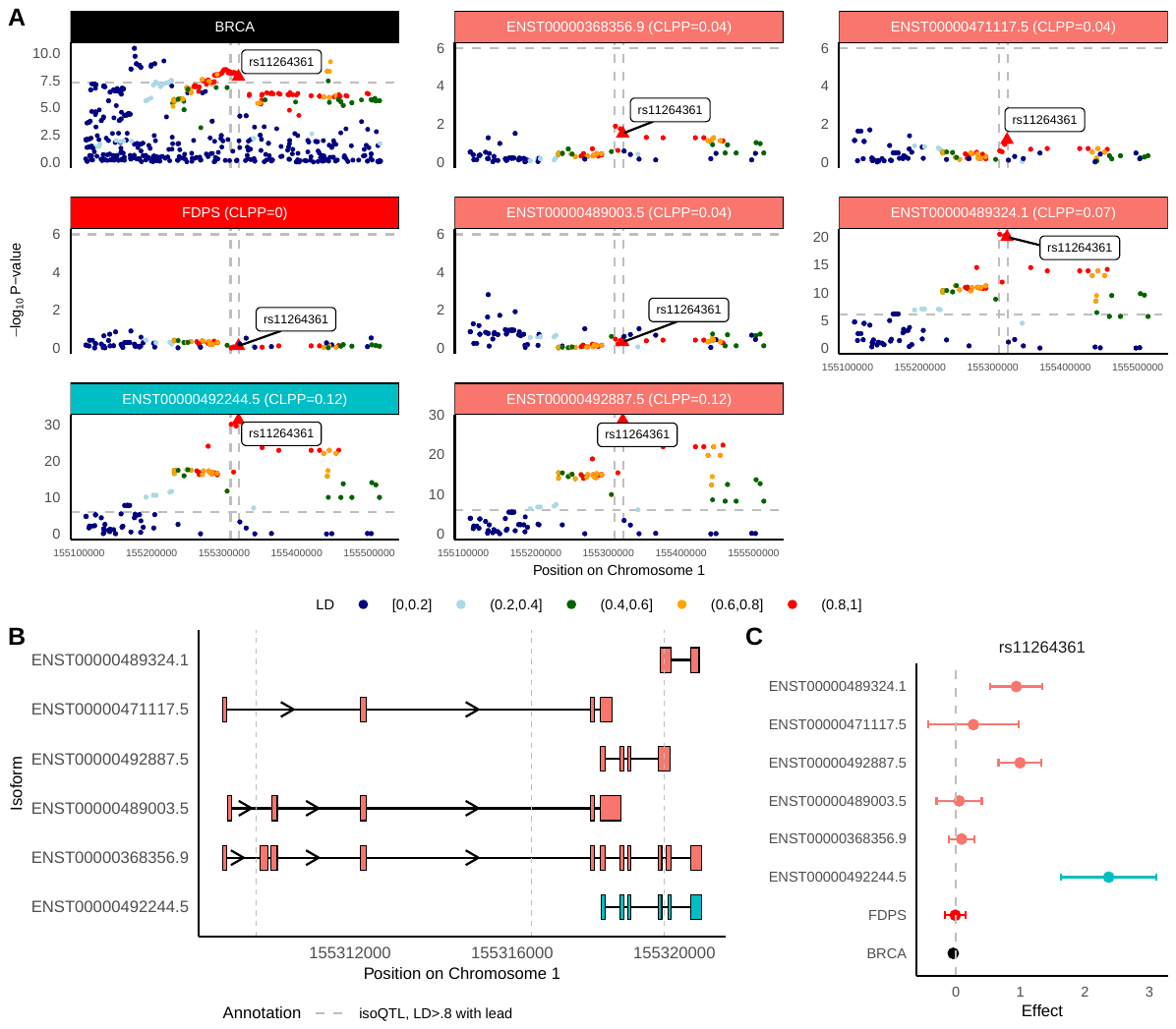
**Figure S24**: *FDPS* *isoforms may mediate breast cancer risk GWAS locus at Chromosome 1q22*. **(A)** Manhattan plot of GWAS effects, *FDPS* gene-eQTLs, and isoform-eQTLs for isoforms of *FDPS*, either prioritized through isoTWAS or with an isoQTL with P < 1e-6. **(B)** Transcript structure of *FDPS*. Vertical lines indicated significant isoQTLs of LD > 0.8 to rs11264361, strongest isoQTL of *FDPS* isoform ENST00000492244.5. **(C)** SNP effect sizes on BRCA risk (black), *FDPS* gene expression (red), expression of isoTWAS-prioritized isoforms (blue), and expression of other isoforms (peach) for rs11264361 (lead isoQTL).

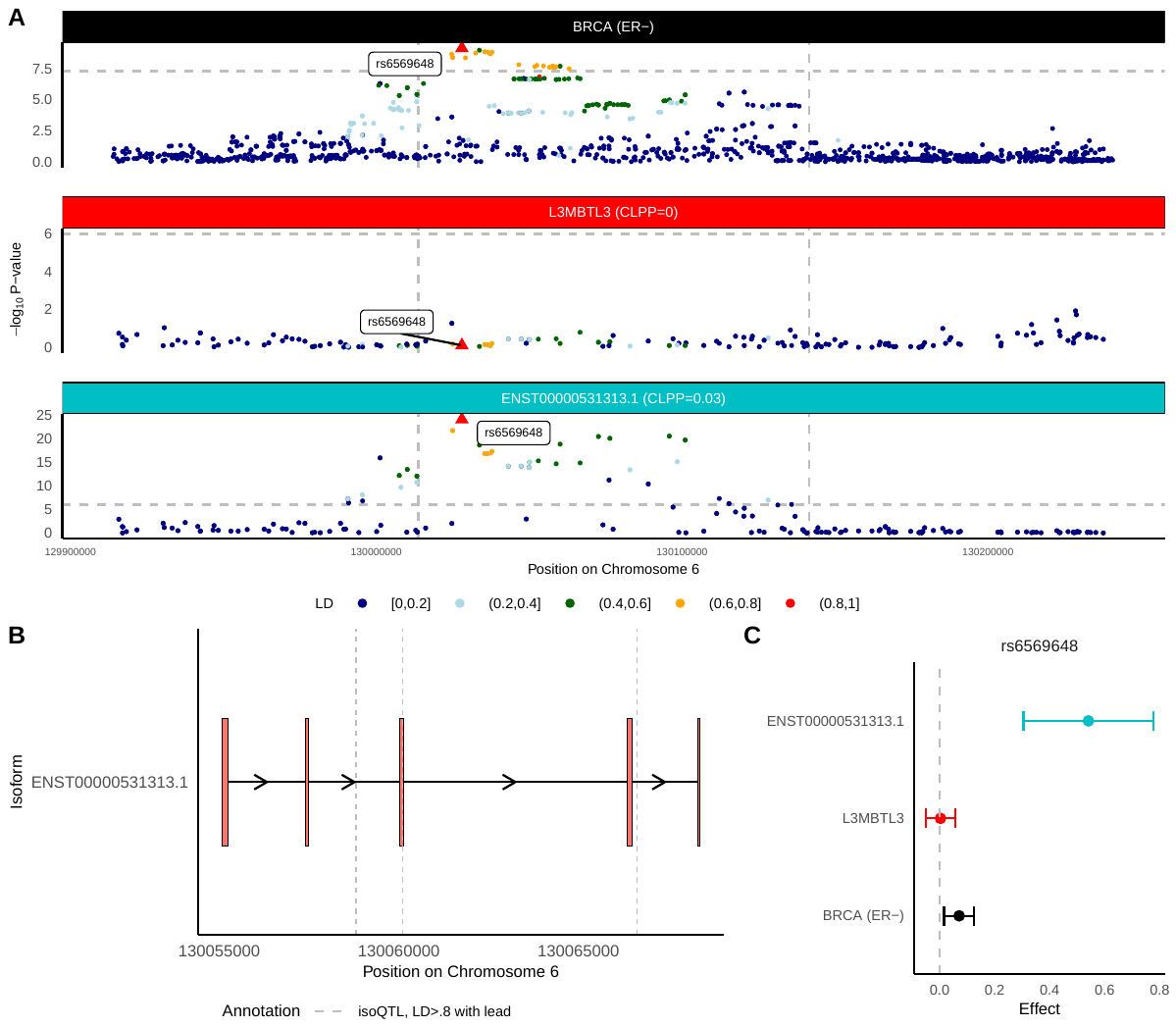
**Figure S25**: *L3MBTL3* *isoform may mediate ER- breast cancer risk GWAS locus at Chromosome 6q23.1*. **(A)** Manhattan plot of GWAS effects, *L3MBTL3* gene-eQTLs, and isoform-eQTLs for isoforms of *L3MBTL3*, either prioritized through isoTWAS or with an isoQTL with P < 1e-6. **(B)** Transcript structure of *L3MBTL3*. Vertical lines indicated significant isoQTLs of LD > 0.8 to rs6569648, strongest isoQTL of *FDPS* isoforms. **(C)** SNP effect sizes on ER- BRCA risk (black), *L3MBTL3* gene expression (red), expression of isoTWAS-prioritized isoforms (blue), and expression of other isoforms (peach) for rs6569648 (lead isoQTL).

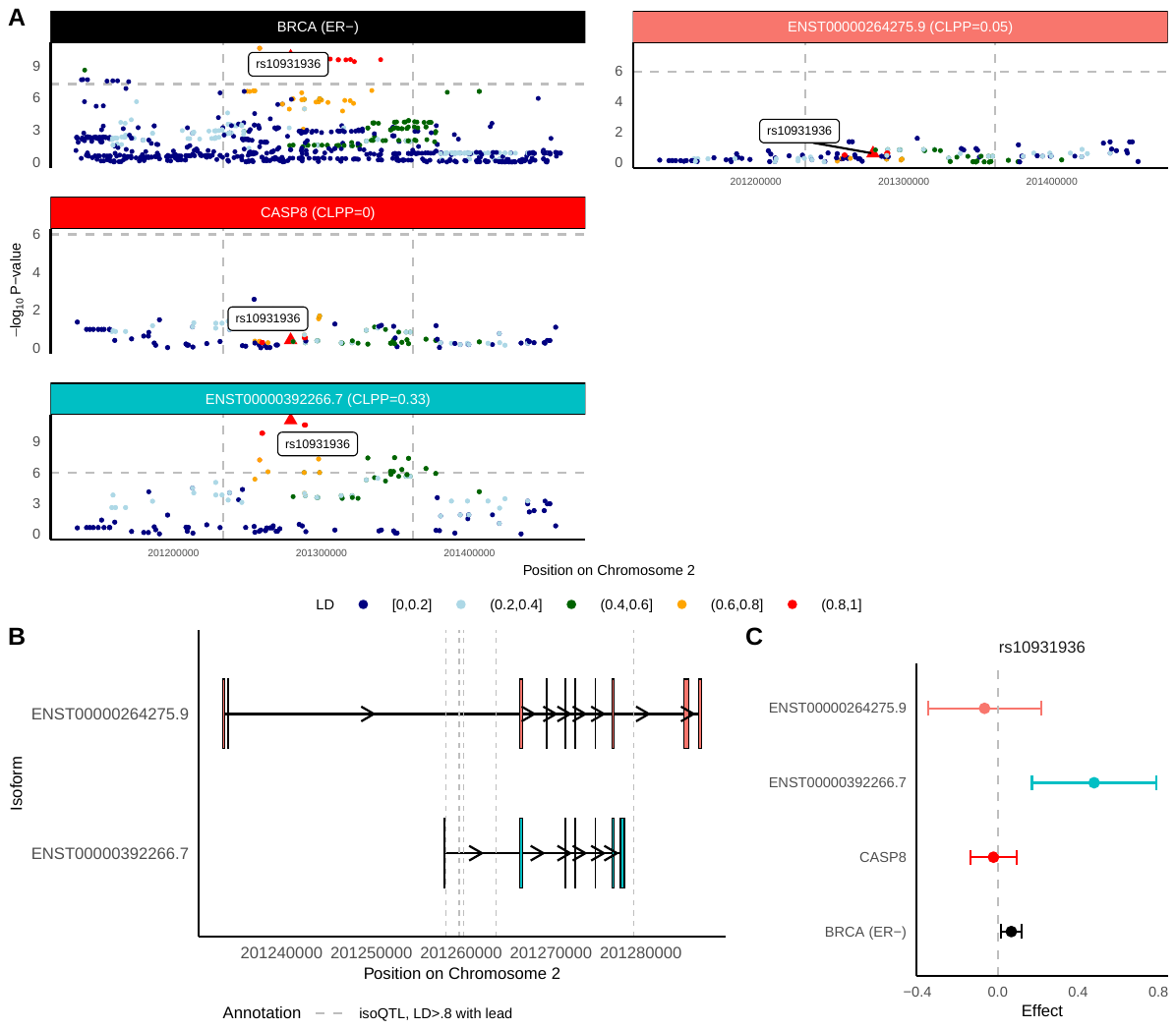
**Figure S26**: *CASP8* *isoforms may mediate ER- breast cancer risk GWAS locus at Chromosome 2q33.1*. **(A)** Manhattan plot of GWAS effects, *CASP8* gene-eQTLs, and isoform-eQTLs for isoforms of *CASP8*, either prioritized through isoTWAS or with an isoQTL with P < 1e-6. **(B)** Transcript structure of *CASP8*. Vertical lines indicated significant isoQTLs of LD > 0.8 to rs10931936, strongest isoQTL of *CASP8* isoforms. **(C)** SNP effect sizes on ER- BRCA risk (black), *CASP8* gene expression (red), expression of isoTWAS-prioritized isoforms (blue), and expression of other isoforms (peach) for rs10931936 (lead isoQTL).

**Figure S27**: *TMBIM1* *isoforms may mediate colorectal cancer risk GWAS locus at Chromosome 2q35*. **(A)** Manhattan plot of GWAS effects, *TMBIM1* gene-eQTLs, and isoform-eQTLs for isoforms of *TMBIM1*, either prioritized through isoTWAS or with an isoQTL with P < 1e-6. **(B)** Transcript structure of *TMBIM1*. Vertical lines indicated significant isoQTLs of LD > 0.8 to rs2382817, strongest isoQTL of *TMBIM1* isoforms. **(C)** SNP effect sizes on CRC risk (black), *TMBIM1* gene expression (red), expression of isoTWAS-prioritized isoforms (blue), and expression of other isoforms (peach) for rs2382817 (lead isoQTL).

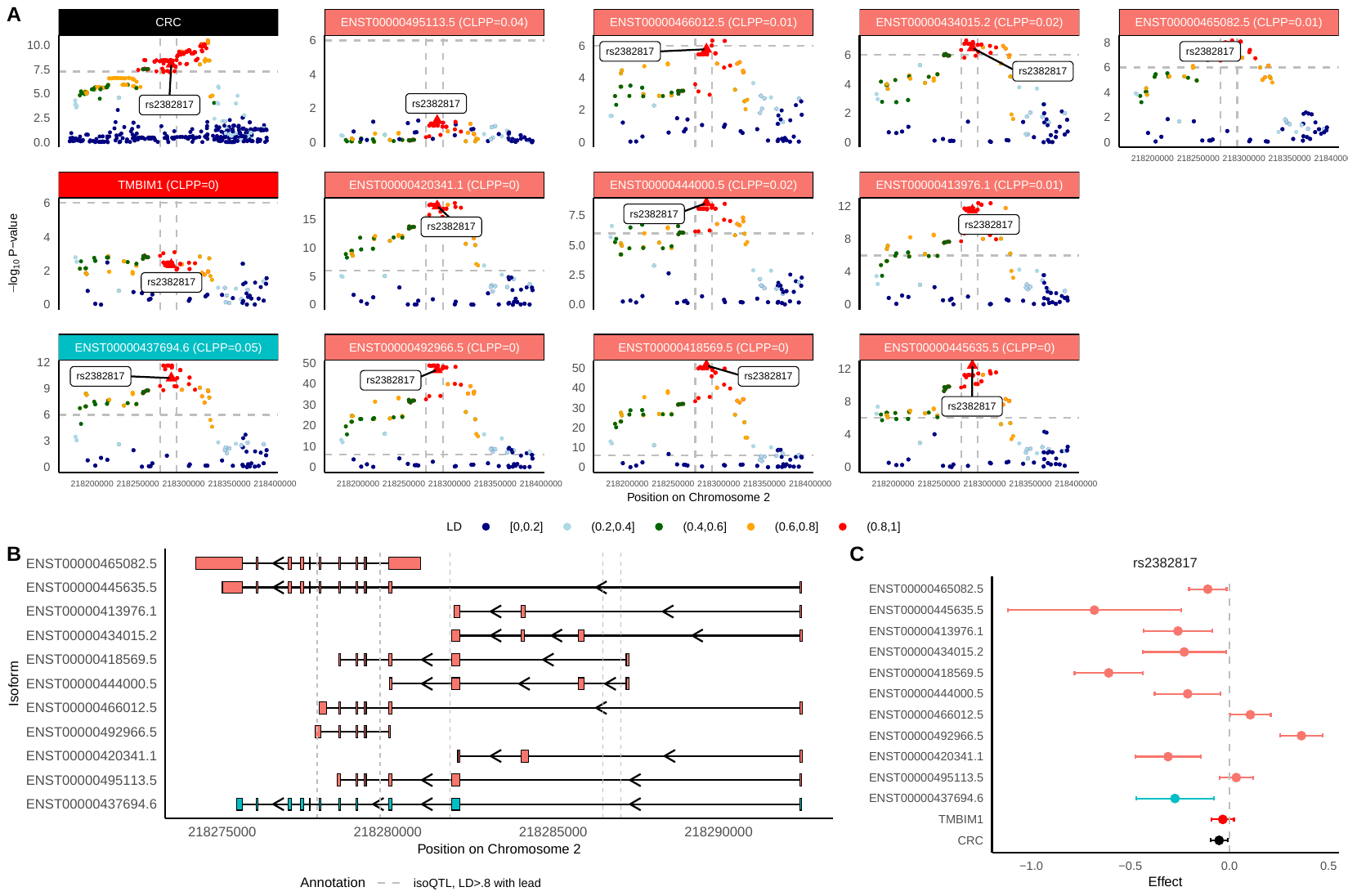

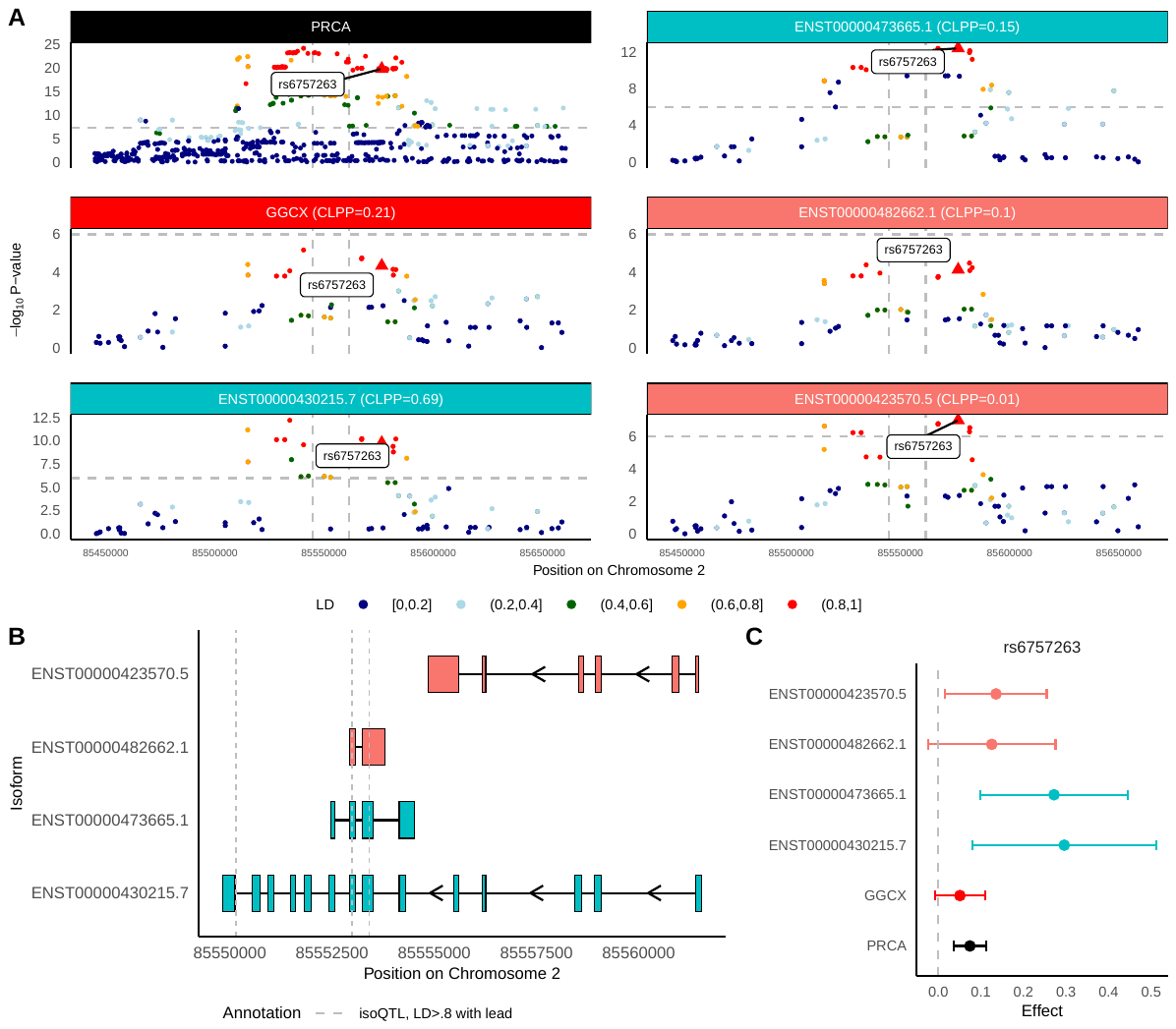

**Figure S28**: *GGCX isoforms may mediate prostate cancer risk GWAS locus at Chromosome 2p11.2*. **(A)** Manhattan plot of GWAS effects, *GGCX* gene-eQTLs, and isoform-eQTLs for isoforms of *GGCX*, either prioritized through isoTWAS or with an isoQTL with P < 1e-6. **(B)** Transcript structure of *GGCX*. Vertical lines indicated significant isoQTLs of LD > 0.8 to rs6757263, strongest isoQTL of *GGCX* isoforms. **(C)** SNP effect sizes on PRCA risk (black), *GGCX* gene expression (red), expression of isoTWAS-prioritized isoforms (blue), and expression of other isoforms (peach) for rs6757263 (lead isoQTL).

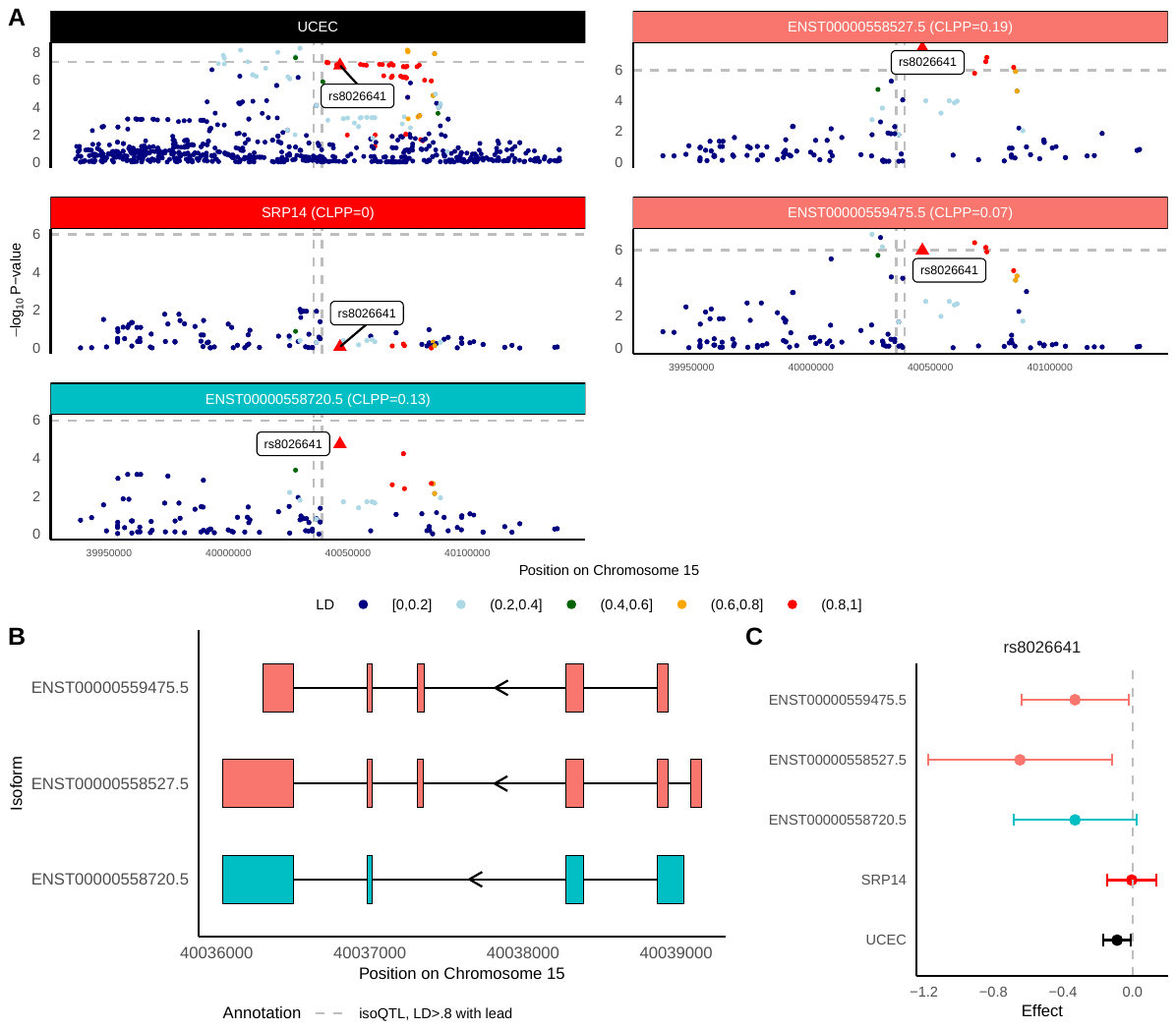
**Figure S29**: *SRP14 isoforms may mediate endometrial cancer risk GWAS locus at Chromosome 15q22*. **(A)** Manhattan plot of GWAS effects, *SRP14* gene-eQTLs, and isoform-eQTLs for isoforms of *SRP14*, either prioritized through isoTWAS or with an isoQTL with P < 1e-6. **(B)** Transcript structure of *SRP14*. **(C)** SNP effect sizes on UCEC risk (black), *SRP14* gene expression (red), expression of isoTWAS-prioritized isoforms (blue), and expression of other isoforms (peach) for rs8026641 (lead isoQTL).

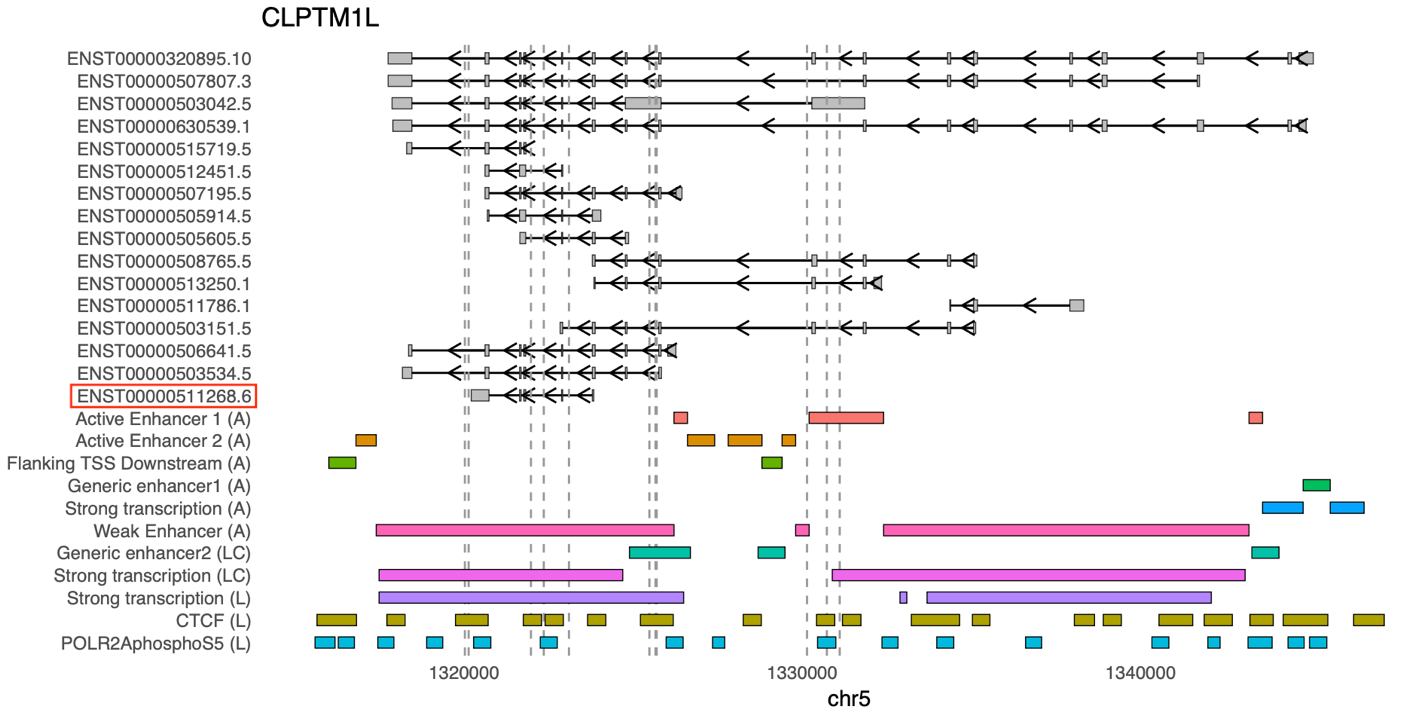

**Figure S30**: *Track plot for CLPTM1L.*(A) means subcutaneous adipose tissue. (LC) mean lung cancer tissue. (L) means lung tissue. Vertical lines indicated significant isoQTLs of LD = 1 to rs414965, strongest isoQTL of *CLPTM1L* isoforms. The prioritized isoform identified by isoTWAS is highlighted in a red box.

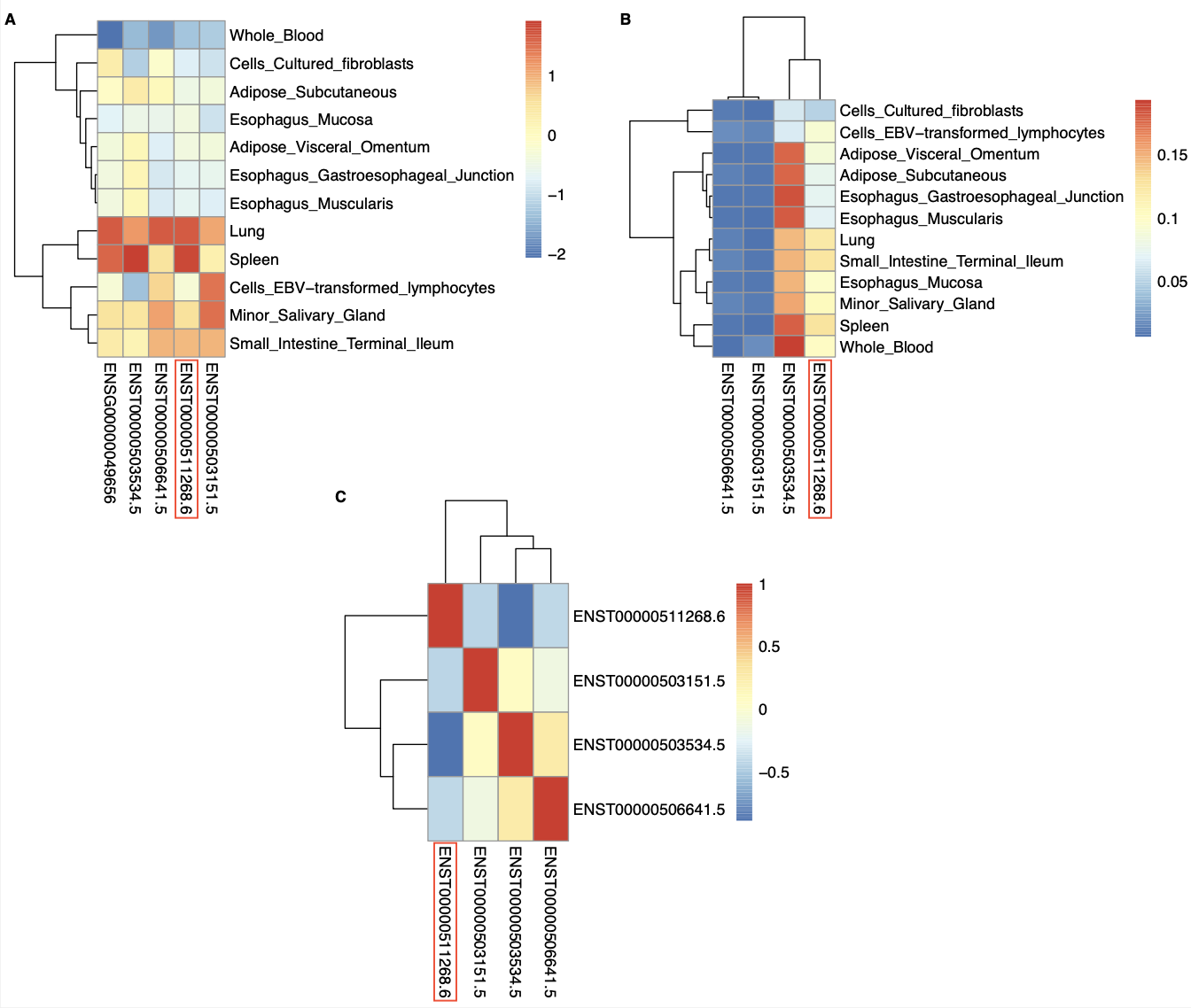

**Figure S31:** *Heatmaps for isoTWAS-identified transcripts in CLPTM1L.* (A) Gene- and isoform-level expression heatmap. (B) Isoform usage heatmap. (C) Correlation heatmap of transcript expression. The prioritized isoform identified by isoTWAS is highlighted in a red box.

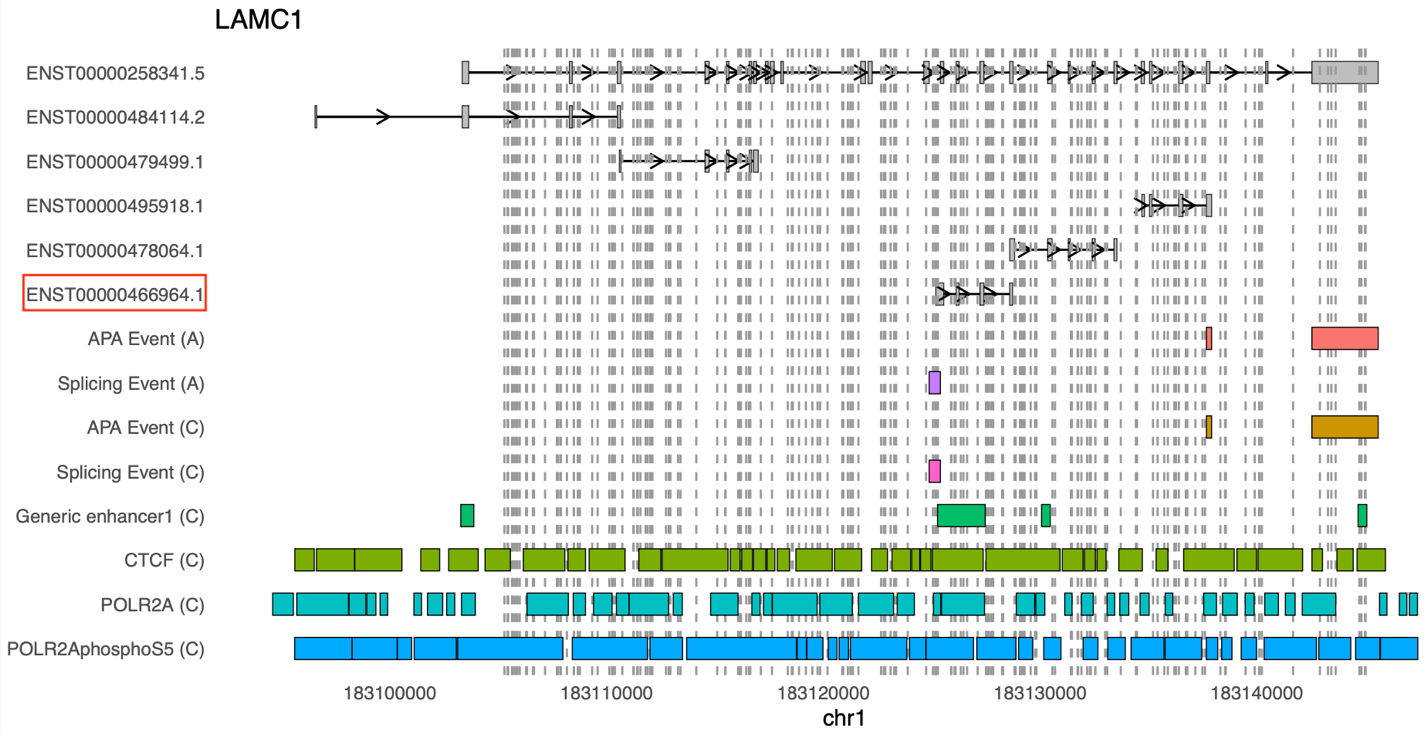

**Figure S32**: *Track plot for LAMC1.* (A) means subcutaneous adipose tissue. (C) mean colon sigmoid tissue. The first intron of transcript ENST00000258341.5 was removed due to its excessive length. Vertical lines indicated significant isoQTLs of LD = 1 to rs20558, strongest isoQTL of *LAMC1* isoforms. The prioritized isoform identified by isoTWAS is highlighted in a red box.

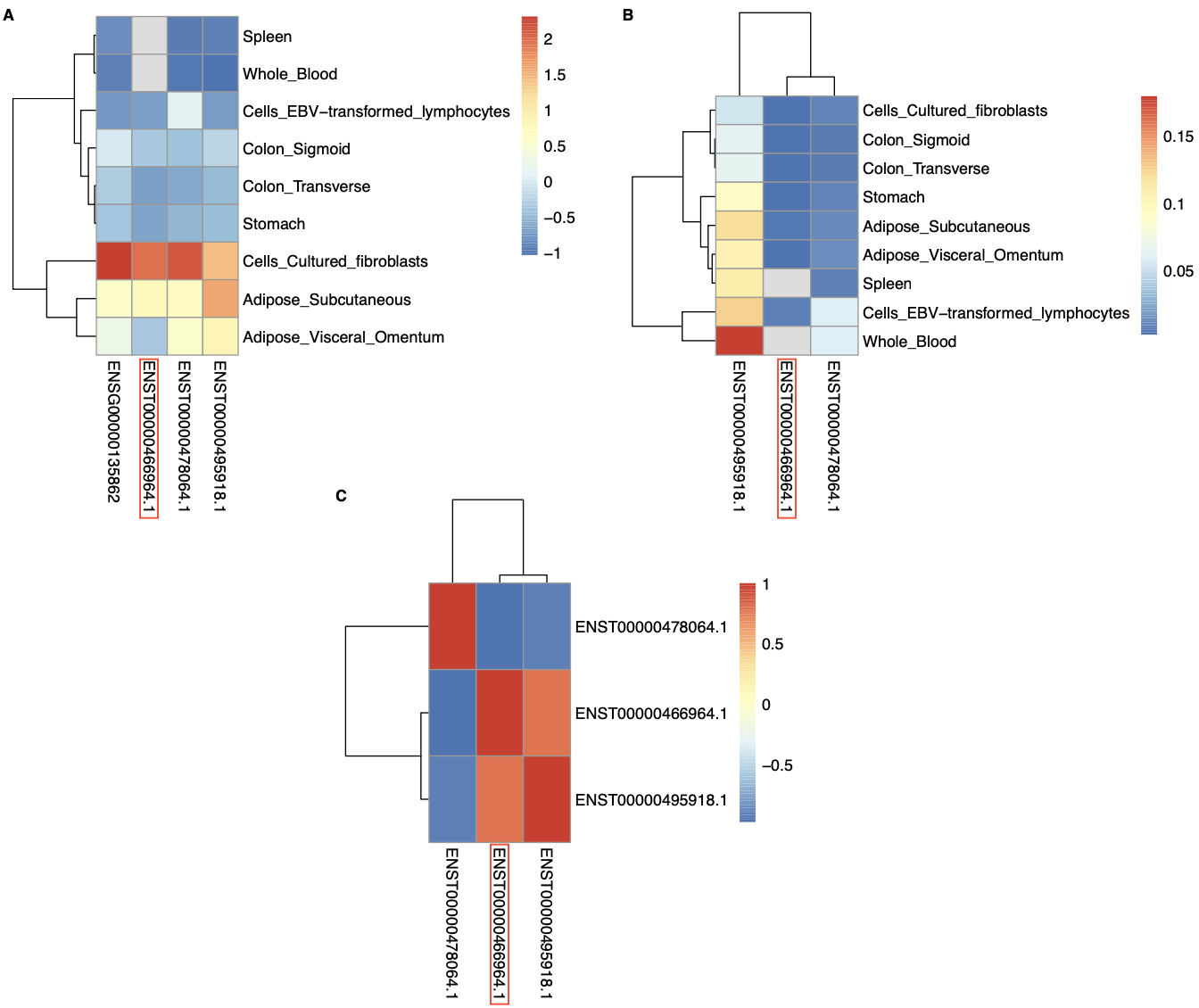

**Figure S33:** *Heatmaps for isoTWAS-identified transcripts in LAMC1.* (A) Gene- and isoform-level expression heatmap. (B) Isoform usage heatmap. (C) Correlation heatmap of transcript expression. The prioritized isoform identified by isoTWAS is highlighted in a red box.

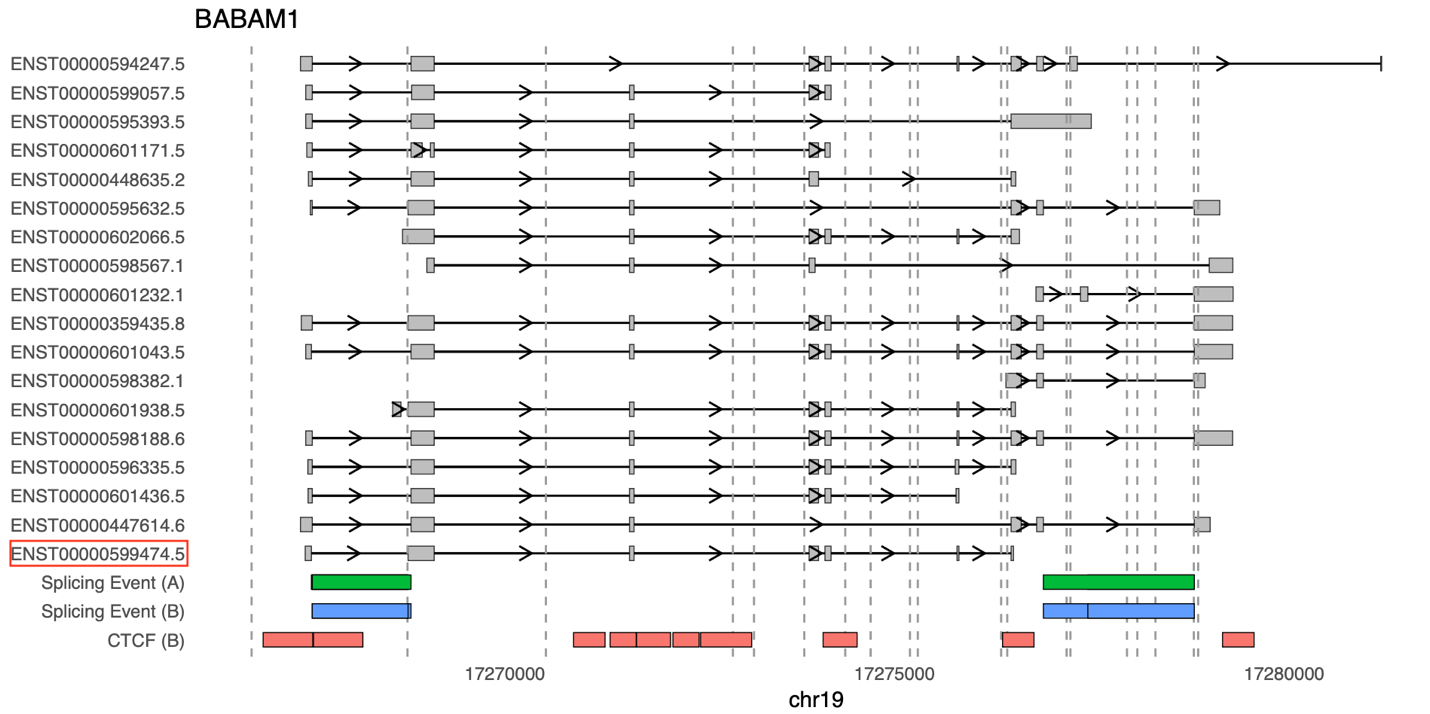

**Figure S34**: *Track plot for BABAM1.* (A) means subcutaneous adipose tissue. (B) mean breast tissue. Vertical lines indicated significant isoQTLs of LD = 1 to rs34084277, strongest isoQTL of *BABAM1* isoforms. The prioritized isoform identified by isoTWAS is highlighted in a red box.

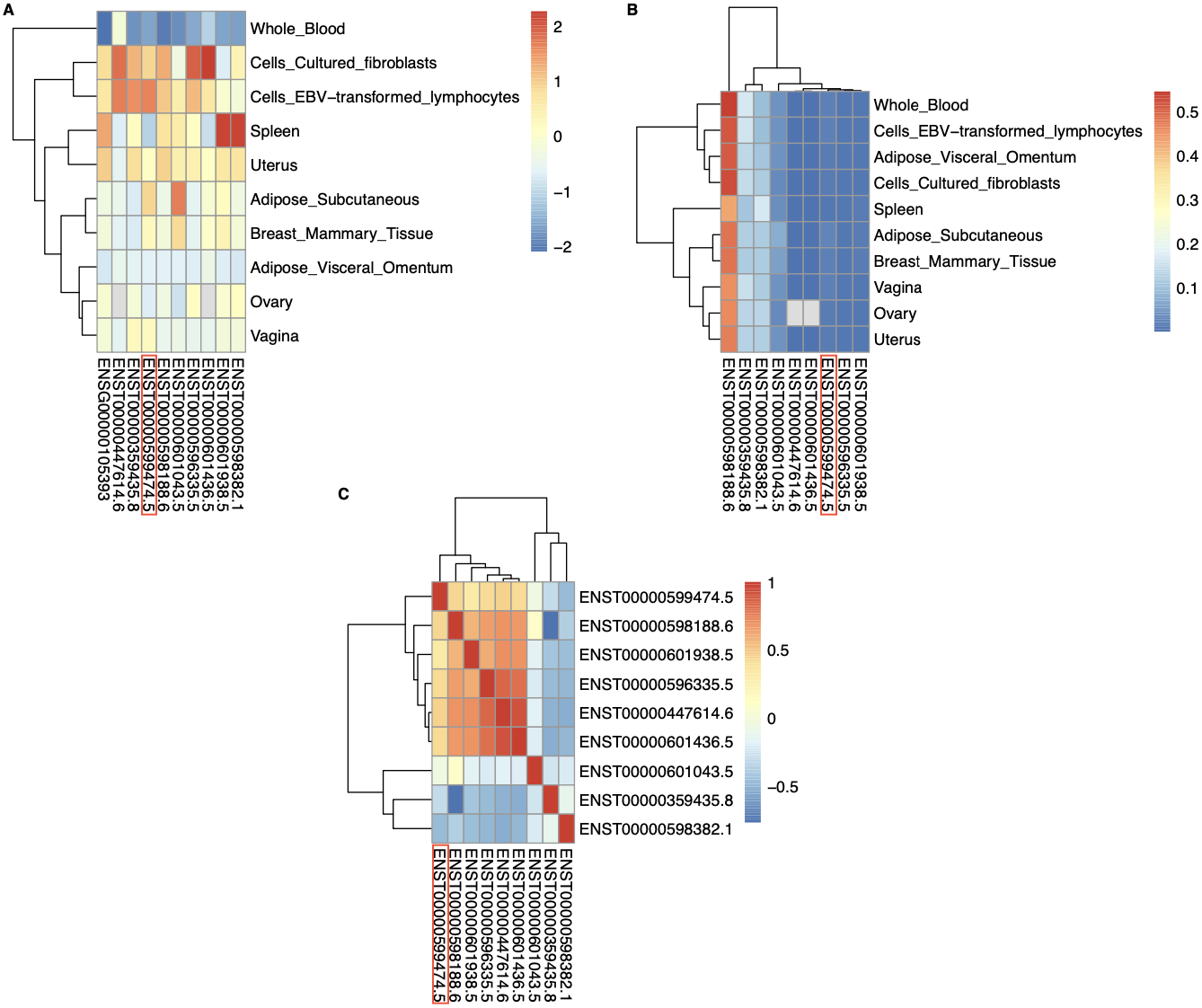

**Figure S35:** *Heatmaps for isoTWAS-identified transcripts in BABAM1.* (A) Gene- and isoform-level expression heatmap. (B) Isoform usage heatmap. (C) Correlation heatmap of transcript expression. The prioritized isoform identified by isoTWAS is highlighted in a red box.

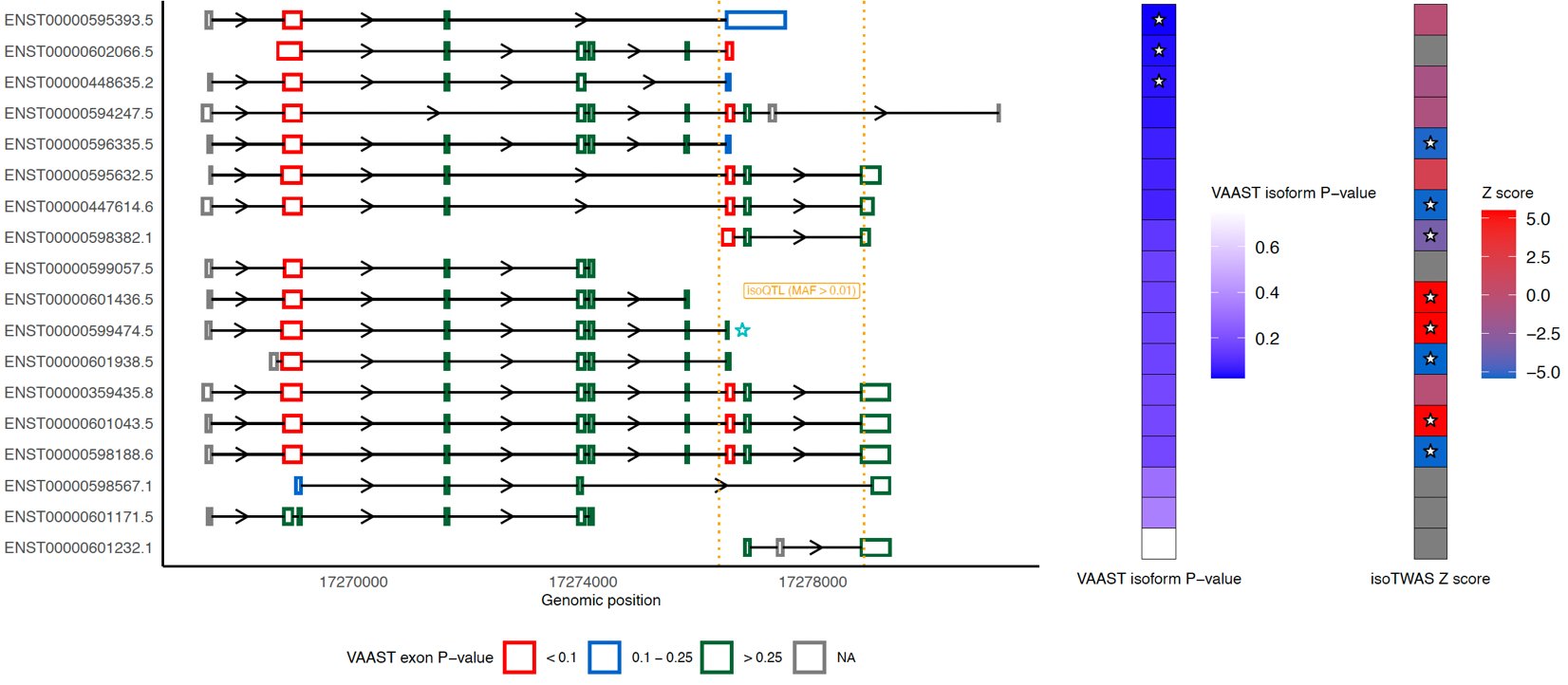

**Figure S36**: *Rare variants analyses may provide further insights into BABAM1.* The figure illustrates the transcript structure of *BABAM1*, with the blue star marking the isoform prioritized from fine-mapping. Orange vertical lines represent significant isoQTLs of the starred isoform with LD > 0.8 relative to rs34084277. The star in the middle heatmap indicates isoforms identified as significant by VAAST (P < 0.05), while the star in the right heatmap denotes isoform-level transcriptome-wide significance (FDR and permutation P < 0.05).

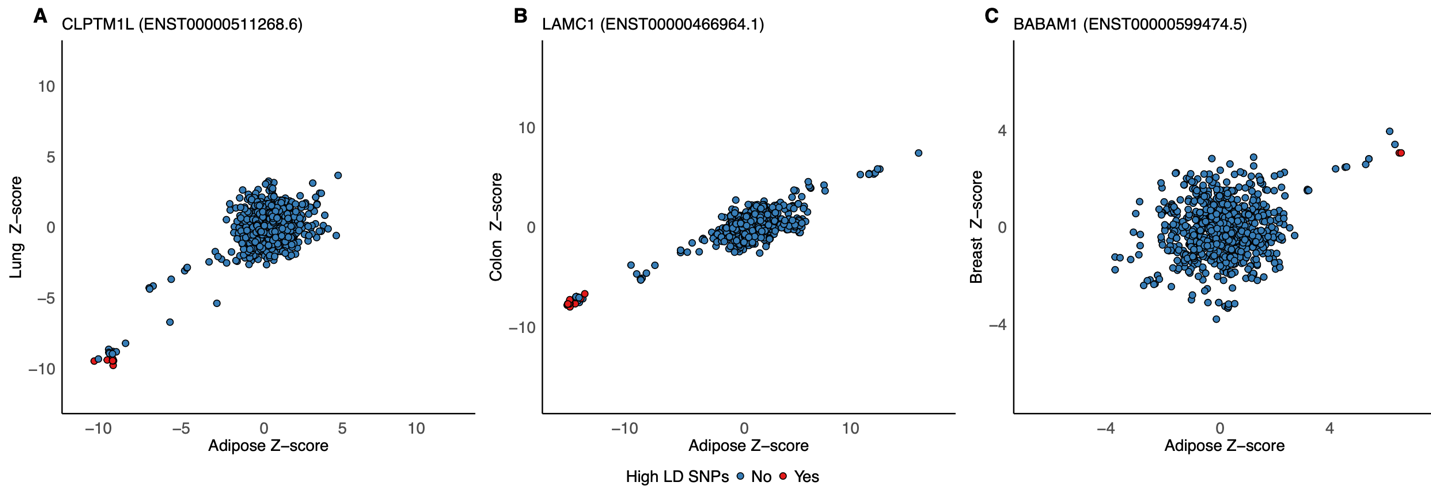

**Figure S37**: *The scatter plots for QTL effect in adipose verse in the tissue of origin.* (A) The scatter plot for *CLPTM1L*, which prioritized isoform is ENST00000511268.6 (B) The scatter plot for *LAMC1*, which prioritized isoform is ENST00000466964.1. (C) The scatter plot for *BABAM1*, which prioritized isoform is ENST00000599474.5. SNPs in high LD (LD > 0.9) with the lead isoQTLs are highlighted in red.

**SUPPLEMENTAL TABLES**

**Table S1**: GWAS sample size and assignments of relevant tissues to cancer indication

**Table S2**: Sample size and tissues for functional genomics reference panels

**Table S3**: isoTWAS and fine-mapping results for genes with FDR-adjusted P < 0.05 and permutation P < 0.05 across 12 cancer indications

**Table S4**: TWAS and fine-mapping results for genes with FDR-adjusted P < 0.05 and permutation P < 0.05 across 12 cancer indications

**Table S5**: Summary of gene-level associations across isoTWAS and TWAS

**Table S6**: Key biological pathways that were enriched for cancer-specific sets of isoTWAS-prioritized genes

**Table S7**: Number of GWAS loci tagged by each method within 0.5 Mb

**Table S8**: isoQTL and eQTL colocalization results

**Table S9**: Estimation of expression-mediated SNP heritability by MESC

**Table S10**: ENCODE and ROADMAP Data Accessions

**Table S11:** SpliceAI Look-Up Results: Variants with Moderate or Higher Predicted Score

**Table S12**: Transcript-based p-values from VAAST in rare variant analysis

**Table S13**: Exon-specific p-values from VAAST in rare variant analysis

**SUPPLEMENTAL DATA**

**Data S1:** Raw isoTWAS results with nominal P-values, screening gene-level P-values, confirmation P-values, and permutation test P-values

**Data S2**: Raw TWAS results with nominal P-values, permutation P-values, and FDR-adjusted P-values

**Data S3**: Gene- and isoform-level QTL summary statistics and colocalization results
